# High-contrast in-vivo imaging of tau pathologies in Alzheimer’s and non-Alzheimer’s disease tauopathies

**DOI:** 10.1101/2020.03.05.20028407

**Authors:** Kenji Tagai, Maiko Ono, Manabu Kubota, Soichiro Kitamura, Keisuke Takahata, Chie Seki, Yuhei Takado, Hitoshi Shinotoh, Yasunori Sano, Kiwamu Matsuoka, Hiroyuki Takuwa, Masafumi Shimojo, Manami Takahashi, Kazunori Kawamura, Tatsuya Kikuchi, Maki Okada, Haruhiko Akiyama, Hisaomi Suzuki, Mitsumoto Onaya, Takahiro Takeda, Kimihito Arai, Nobutaka Arai, Nobuyuki Araki, Yuko Saito, Yasuyuki Kimura, Masanori Ichise, Yutaka Tomita, Ming-Rong Zhang, Tetsuya Suhara, Masahiro Shigeta, Naruhiko Sahara, Makoto Higuchi, Hitoshi Shimada

**Author notes:** These authors contributed equally to this work (Kenji Tagai, Maiko Ono, Manabu Kubota). Correspondence (M.Higuchi).

## Abstract

A panel of radiochemicals has enabled *in-vivo* positron emission tomography (PET) of tau pathologies in Alzheimer’s disease (AD), while sensitive detection of frontotemporal lobar degeneration (FTLD) tau inclusions has been unsuccessful. Here, we generated an imaging probe, PM-PBB3, for capturing diverse tau deposits. *In-vitro* assays demonstrated the reactivity of this compound with tau pathologies in AD and FTLD. We could also utilize PM-PBB3 for optical/PET imaging of a living murine tauopathy model. A subsequent clinical PET study revealed increased binding of ^18^F-PM-PBB3 in diseased patients, reflecting cortical-dominant AD and subcortical-dominant PSP tau topologies. Notably, the *in-vivo* reactivity of ^18^F-PM-PBB3 with FTLD tau inclusion was strongly supported by neuropathological examinations of autopsied and biopsied brains derived from Pick’s disease, PSP and corticobasal degeneration patients who underwent PET scans. Finally, visual inspection of ^18^F-PM-PBB3-PET images was indicated to facilitate individually based identification of diverse clinical phenotypes of FTLD on the neuropathological basis.

## INTRODUCTION

The vast majority of age-related neurodegenerative diseases are characterized as protein conformational disorders, involving self-assemblies of misfolded proteins into fibrillary aggregates (Soto and Pritzkow, 2018; Walker and Jucker, 2015). Among these pathogenic proteins, the fibrillogenesis of microtubule-associated protein tau occurs as a hallmark pathological change in diverse illnesses referred to as tauopathies, and it is mechanistically linked to the neurodegenerative processes in these disorders (Iqbal et al., 2016; Spillantini and Goedert, 2013). Tau in the central nervous system is composed of six isoforms, which are classified into three- and four-repeat species according to the number of repeat domains (Buee et al., 2000). Alzheimer’s disease (AD) and AD-type primary age-related tauopathy (PART) are characterized by tau pathologies formed by all isoforms, while a significant subset of frontotemporal lobar degeneration (FTLD) syndromes is neuropathologically unfolded by exclusive fibrillization of either three- or four-repeat tau isoforms (Buee et al., 2000; Lee et al., 2001). The differences in the isoform composition among these tauopathies lead to diversities in the conformation and ultrastructures of tau fibrils as revealed by recent cryo-electron microscopic assays (Falcon et al., 2018; Fitzpatrick et al., 2017).

The distinct tau conformers are likely to determine subcellular, cellular, and regional localization of tau deposits in a disease-specific fashion, provoking characteristic symptoms associated with deteriorations of affected neurons and neural circuits (Forrest et al., 2019). In line with this mechanism, there exist clear distinctions among neuropathological features of AD/PART and major tau-positive FTLD disorders, including three-repeat tauopathies represented by Pick’s disease (PiD) and four-repeat tauopathies exemplified by progressive supranuclear palsy (PSP) and corticobasal degeneration (CBD) (Lee et al., 2001). Meanwhile, substantial overlaps have been noted among symptomatic phenotypes derived from these pathologies, impeding the differentiation of clinical syndromes by estimation of underlying pathological alterations (Rabinovici and Miller, 2010; Williams and Lees, 2009; Zhang et al., 2020).

*In-vivo* imaging technologies such as positron emission tomography (PET) with specific radioligands for amyloid-beta and tau fibrils have enabled visualization of AD-type neuropathologies in living subjects, facilitating diagnosis and staging of AD dementia and its prodrome. The tau PET probes available for these clinical assays are classified into three chemotypes consisting of ^18^F-labeled THK5351 (Harada et al., 2016), ^18^F-labeled flortaucipir (Chien et al., 2014), and ^11^C-labeled PBB3 (Maruyama et al., 2013; Shimada et al., 2017) series originating from nonclinical prototypes BF-158/BF-170 (Okamura et al., 2005), BF-126 (Okamura et al., 2005), and styryl 7 (PBB5) (Maruyama et al., 2013), respectively. Unlike for AD tau lesions, high-contrast PET detection of three- and four-repeat tau deposits in FTLD patients has been unsuccessful, as tau-related radiosignals yielded by ^18^F-flortaucipir and ^11^C-PBB3 in PSP and CBD cases were less than 20% of the corresponding signals in patients with advanced AD (Endo et al., 2019; Schonhaut et al., 2017). ^18^F-THK5351 was reported to illuminate brain areas putatively enriched with PSP and CBD tau inclusions (Brendel et al., 2017; Kikuchi et al., 2016), but those observations were attributed to the cross-reactivity of this compound with monoamine oxidase B (MAO-B), which is upregulated in reactive astrocytes (Harada et al., 2017; Ng et al., 2017). In addition, most ‘second-generation’ tau PET probes are analogs of ^18^F-fluotaucipir and are not overtly more reactive with non-AD tau assemblies than ^18^F-fluotaucipir and ^11^C-PBB3 (Aguero et al., 2019; Honer et al., 2018; Matthias Brendel, 2019).

^11^C-PBB3 was originally designed to capture tau fibrils in a wide range of tauopathies (Maruyama et al., 2013) and was demonstrated to react with three- and four-repeat tau aggregates in human brain tissues with a higher binding potential than ^18^F-flortaucipir (Ono et al., 2017). However, rapid conversion of ^11^C-PBB3 into a metabolite resulted in a relatively low entry of the unmetabolized compound into the brain (Hashimoto et al., 2014; Kimura et al., 2015; Maruyama et al., 2013), hampering a sensitive recognition of fibrillary aggregates in FTLD tauopathies that are less abundant than AD tau deposits. To overcome this technical issue, in the current work we modified the chemical structure of PBB3 to generate a chemical with a relatively high metabolic stability, aiming at unambiguous investigations of tau fibril density and extent in each of the individuals with AD and FTLD. The new compound, PM-PBB3 (propanol modification of PBB3), was also fluorinated in consideration of advantages of an ^18^F-labeled probe over ^11^C-radiochemicals for broader availability and higher PET scan throughput. Nonclinical assays revealed the capability of PM-PBB3 for high-sensitivity illumination of tau pathologies in a murine model bimodally by *in-vivo* optical and PET imaging from single-cell to brain-wide scales, potentially serving for the discovery of candidate therapeutics counteracting the neurodegenerative tau pathogenesis. Subsequent applications of ^18^F-PM-PBB3 to clinical PET assays, along with neuropathological data obtained from scanned subjects, demonstrated appropriate kinetic and binding profiles of this probe for personalized evaluations of tau depositions in AD and various FTLD syndromes.

## RESULTS

### *In-vitro* binding of PM-PBB3 to AD- and FTLD-type tau aggregates

The original compound, PBB3 (Figure 1a), was found to be promptly conjugated with sulfate at a hydroxy moiety following systemic injection (Hashimoto et al., 2014). To suppress this metabolic conversion, we substituted this substructure with the fluoroisopropanol group, resulting in the generation of PM-PBB3 (Figure 1a). This modification also allowed ^18^F radiolabeling of the probe using a tosylate precursor (Figure S1). Similar to PBB3 (Maruyama et al., 2013; Ono et al., 2017), PM-PBB3 is self-fluorescent, and its reactivity with pathological tau fibrils is assessable by fluorescence labeling of brain sections derived from tauopathy patients. Triple staining of brain slices with PM-PBB3, antibody against phosphorylated tau (AT8), and Gallyas-Braak silver impregnation (GB) demonstrated binding of PM-PBB3 to neurofibrillary tangles (NFT), neuropil threads, and dystrophic neurites encompassing neuritic plaques in AD hippocampal formation, which were composed of six tau isoforms (Figure 1b). Furthermore, PM-PBB3 conspicuously illuminated Pick bodies constituted of three-repeat tau isoforms in PiD frontal cortex and four-repeat tau lesions such as tufted astrocytes in PSP striatum, astrocytic plaques in CBD striatum, and coiled bodies and argyrophilic grains and threads in these tissues (Figure 1b).

**Figure 1.**
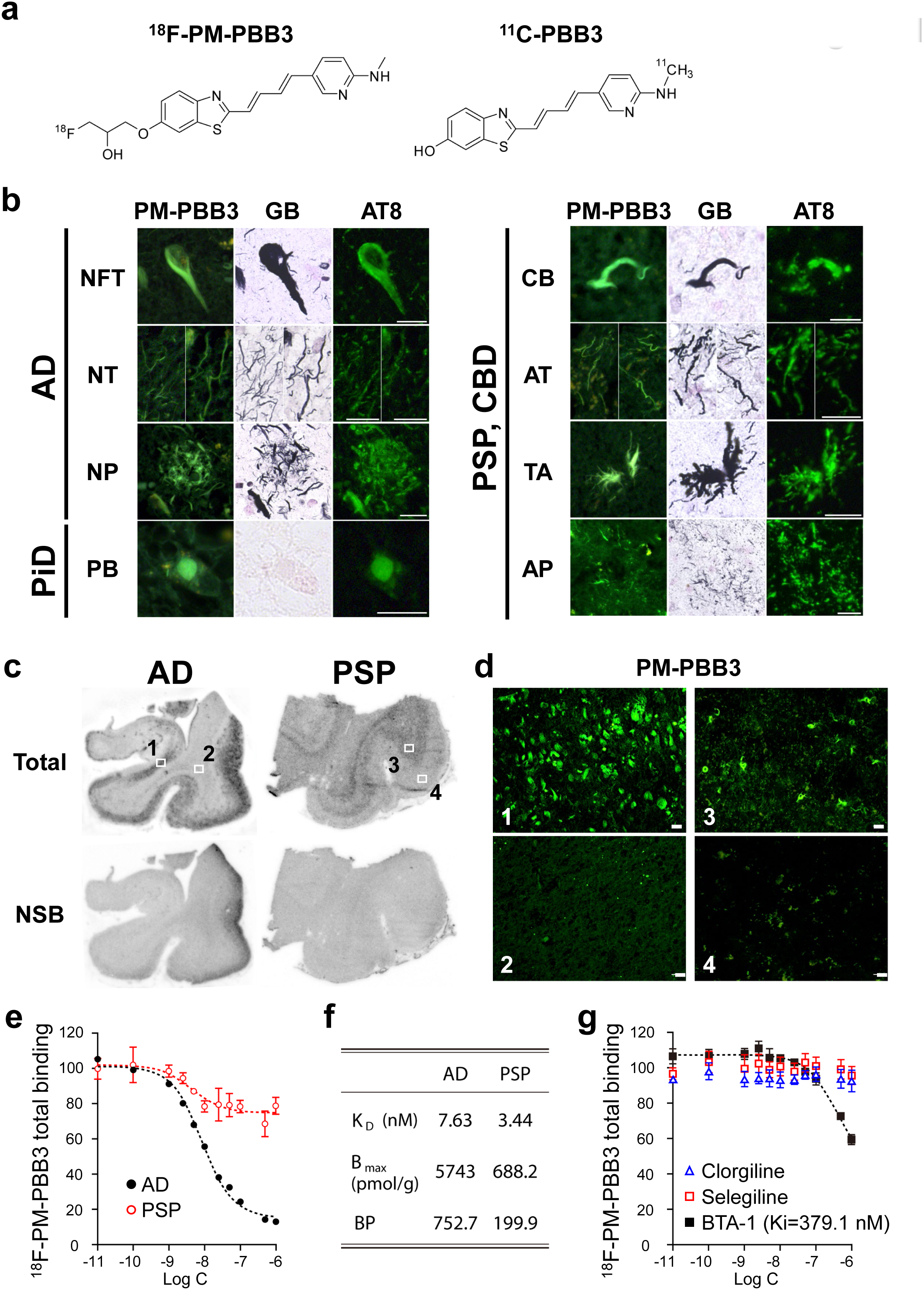
*In vitro* binding of PM-PBB3 to tau lesions in AD, PiD, PSP and CBD. (**a**) Chemical structural formulae of ^18^F-PM-PBB3 (left) and ^11^C-PBB3 (right). (**b**) Triple staining of tau lesions in the hippocampal formation of an AD patient (left composite), the frontal cortex of a PiD patient (left composite) and the caudate/putamen of PSP and CBD patients (right composite) with 25 μM of non-radiolabel ed PM-PBB3, GB, and AT8. NFT, neuropil threads (NT) and dystrophic neurites encompassing a neuritic plaque (NP) in AD brain sections, pick body (PB) in PiD brain section, and coiled body (CB), argyrophilic threads (AT), tufted astrocyte (TA) and astrocytic plaque (AP) in PSP/CBD brain sections are clearly labeled with PM-PBB3. Scale bars, 20 μm. (**c**) Autoradiographic labeling of AD brain sections including the hippocampal formation and inferior temporal cortex (left) and a PSP motor cortex section (right) with 5 nM of ^18^F-PM-PBB3 in the absence (top, total binding) and presence (bottom, non-specific binding: NSB) of 100 μM of non-radiolabeled PBB5, an analog of PM-PBB3. (**d**) Photomicrographs of fluorescence staining with PM-PBB3 in areas indicated by squares in **c**. In line with autoradiographic data, NFT in the AD subiculum (1), and coiled bodies and tufted astrocytes in middle gray matter layers of the PSP motor cortex (3) were intensely labeled with 25 μM of non-radiolabeled PM-PBB3, in contrast with the lack of overt fluorescence signals in the white matter of the AD temporal cortex (2), and superficial gray matter layers of the PSP motor cortex (4). Scale bars, 20 μm. (**e**) Total (specific + non-specific) bindings of 1 nM of ^18^F-PM-PBB3 in AD frontal cortex (closed circles) and PSP motor cortex (open circles) samples were homologously blocked by non-radiolabeled PM-PBB3 with varying concentrations, and a one-site binding model was employed for describing the inhibition plots. Data are mean values ± SD in four samples and are expressed as % of the averaged total binding. (**f**) Binding parameters for ^18^F-PM-PBB3 determined by non-linear fitting of a one-site homologous blockade model to data shown in **e**. (**g**) Inhibition of total binding of 1 nM of [^18^F]PM-PBB3 by clorgiline (MAO-A inhibitor, blue triangle), selegiline (MAO-B inhibitor, red square) and BTA-1 (analog of PiB, black square) in an AD frontal cortex sample. A part of ^18^F-PM-PBB3 total binding was heterologously blocked by BTA-1 with relatively large Ki (379.1 nM). Total binding of ^18^F-PM-PBB3 was not inhibited by clorgiline and selegiline at varying concentrations, while Ki for clorgiline and selegiline was not determined due to failures of the model fitting. Data are mean values ± SD in four samples and are expressed as % of the averaged total binding.

We then radiosynthesized ^18^F-PM-PBB3 and examined its *in-vitro* binding characteristics. Autoradiography of tissue sections demonstrated that ^18^F-PM-PBB3 radiosignals were intensely distributed in the anatomical structures enriched with AD and PSP tau fibrils, as exemplified by gray matter of the hippocampal formation and inferior temporal cortex in the AD brain and gray and white matter of the motor cortex in the PSP brain (Figure 1c). Radioligand binding was profoundly abolished by excessive non-radioactive PBB5 (Figure 1c). Localization of the autoradiographic labeling was in line with histological features obtained from the same sections, as abundant NFTs and neuropil threads in the AD subiculum (area 1), and coiled bodies and tufted astrocytes in middle gray matter layers of the PSP motor cortex (area 3) were captured by nonradiolabeled PM-PBB3 (areas 1, 3 in Figure 1d). In contrast, the lack of overt autoradiographic radioligand binding spatially agreed with minimal PM-PBB3 fluorescence in white matter of the AD temporal cortex, and in superficial gray matter layers of the PSP motor cortex (areas 2, 4 in Figure 1d).

We also quantified the affinity of ^18^F-PM-PBB3 for tau aggregates in homogenized AD frontal cortical and PSP motor cortical tissues. Radioligand binding in these tissues was homologously blocked by non-radiolabeled PM-PBB3 in a concentration-dependent fashion (Figure 1e), indicating binding saturability. ^18^F-PM-PBB3 displayed high-affinity, high-capacity binding in AD homogenates [dissociation constant (K_D_), 7.63 nM; concentration of binding components (B_max_), 5743 pmol/g; binding potential (BP = B_max_ / K_D_), 752.7] (Figure 1f). The radioligand bound in PSP tissues with lower capacity but higher affinity than in AD tissues (K_D_, 3.44 nM; B_max_, 688.2 pmol/g; BP, 199.9). The BP for [^18^F]PM-PBB3 in PSP homogenates was 1.6 times higher than the value for [^11^C]PBB3 in the same samples (Ono et al., 2017). The binding of ^18^F-PM-PBB3 in AD homogenates was partially and heterologously blocked by BTA-1, which is a Pittsburgh Compound-B (PiB) analog and binds to Aβ aggregates with high affinity, with a large inhibition constant (Ki) value (379.1 nM) (Figure 1g), suggesting that ^18^F-PM-PBB3 is incapable of sensitively capturing Aβ deposits in AD homogenates (Klunk et al., 2001; Ni et al., 2018). Moreover, the heterologous blockade by BTA-1 is likely to stem primarily from its low-affinity binding to AD tau fibrils. Notably, minimal displacement of ^18^F-PM-PBB3 binding was observed in the presence of the monoamine oxidase A (MAO-A) inhibitor clorgiline, or the (MAO-B) inhibitor selegiline, in AD frontal cortex homogenates (Figure 1g), suggesting that ^18^F-PM-PBB3 barely cross-reacts with off-target binding sites on monoamine oxidases, unlike the reported binding of ^18^F-THK5351 and ^18^F-flortaucipir to MAO-B (Harada et al., 2017; Lemoine et al., 2018; Ng et al., 2017) and/or MAO-A (Vermeiren et al., 2017).

### Optical and PET detection of tau deposits in living tauopathy model mice

For assessing *in-vivo* interactions of PM-PBB3 with intracellular tau deposits, we utilized a murine transgenic (Tg) model of tauopathies dubbed rTg4510, which overexpresses a human four-repeat tau isoform with the P301L mutation causative of familial FTLD (Sahara et al., 2014; Santacruz et al., 2005). ^18^F-PM-PBB3 bound to tau fibrils in homogenized forebrain tissues obtained from Tg with high affinity (K_D_, 4.7 nM), while there was no homologously displaceable radioligand binding in non-transgenic (nTg) forebrain homogenates (Figure 2a). *Ex-vivo* autoradiography of brain tissues collected from mice at 30 min after intravenous ^18^F-PM-PBB3 injection demonstrated accumulations of the radioligand in the Tg forebrain harboring neuronal tau inclusions (Figure 2b). Conversely, there was no noticeable increase of ^18^F-PM-PBB3 retentions in the nTg forebrain (Figure 2b). In addition, the radioligand accumulation was minimal in the Tg and nTg cerebellum, which was devoid of tau pathologies (Figure 2b). Triple staining of brain sections used for *ex-vivo* autoradiography with PM-PBB3 fluorescence, GB, and AT8 illustrated strong binding of PM-PBB3 to intracellular tau aggregates in the hippocampus and neocortex of a Tg mouse (Figure 2c).

**Figure 2.**
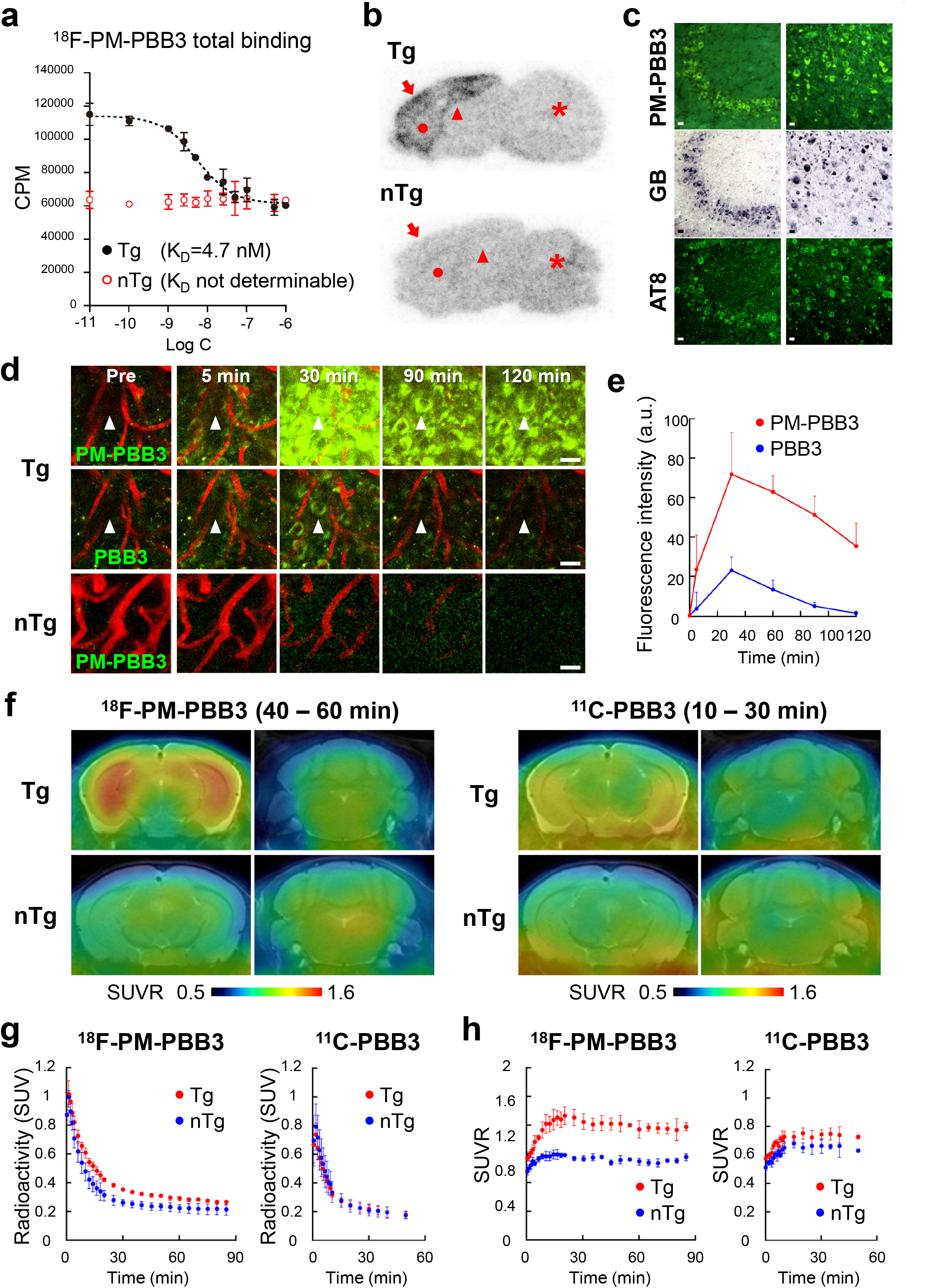
*In vivo* performances of PM-PBB3 as a multimodal probe for optical and PET imaging of tau aggregates in a transgenic mouse model. (**a**) Total (specific + non-specific) bindings of 1 nM of ^18^F-PM-PBB3 in forebrain samples obtained from rTg4510 transgenic (Tg, closed circles) and non-transgenic (nTg, open circles) mice were homologously blocked by non-radiolabeled PM-PBB3 with varying concentrations, and a one-site binding model was employed for describing the inhibition plots. Data are mean values ± SD in four samples and are expressed as cpm. (**b**) *Ex vivo* autoradiographic labeling with intravenously injected ^18^F-PM-PBB3 in 9.7-month-old Tg (top) and nTg (bottom) control mice. The brains were removed at 30 min after injection and were cut into sagittal slices. Significant accumulation of ^18^F-PM-PBB3 was observed in the hippocampus (arrowhead), neocortex (arrow) and striatum (circle) of a Tg mouse, but not in the cerebellum (asterisk). On the other hand, no significant accumulation of ^18^F-PM-PBB3 showed in these brain areas of a nTg mouse. (**c**) Postmortem triple staining of Tg brain sections used in *ex vivo* autoradiographic experiment with 25 μM of non-radiolabeled PM-PBB3, GB and AT8. Numerous intracellular deposits in the hippocampal (left) and neocortical (right) areas corresponding to portions indicated by arrowhead and arrow, respectively, in Panel **b** were strongly labeled with PM-PBB3, GB and AT8. Scale bars, 20 μm. (**d**) *In vivo* two-photon laser microscopic images showing a maximum intensity projection of fluorescence signals in a 3D volume of the somatosensory cortex of 8-month-old Tg (top and middle) and nTg (bottom) mice. Cerebral blood vessels were labeled in red with intraperitoneally administered sulforhodamine 101, and tau aggregates in the Tg mouse were illuminated in green with intravenously injected non-radiolabeled PM-PBB3 (top) or PBB3 (middle), in contrast to minimal retentions of PM-PBB3 in the nTg mouse brain (bottom). Images were acquired before (Pre) and 5, 30, 90 and 120 min after the tracer administration. Arrowheads denote the same tau inclusion detected by PM-PBB3 and PBB3. Scale bars, 20 μm. (**e**) Chronological changes of fluorescence signals derived from PM-PBB3 (red circles) and PBB3 (blue circles) in identical neurons bearing tau aggregate in the somatosensory cortex of Tg mouse over 120 min after the tracer administration in *in vivo* two-photon microscopic imaging. Fluorescence signal intensities normalized accordingly to the background signals were expressed as arbitrary units (a. u.), and data are mean values ± SD in 10 neurons. (**f**) Coronal brain images of 9-month-old Tg (top) and nTg (bottom) mice acquired by averaging dynamic PET data at 40 – 60 and 10 – 30 min after intravenous administration of ^18^F-PM-PBB3 (left composite) and ^11^C-PBB3 (right composite), respectively. Brain volume data were sectioned at 3 mm (left column in each composite) and 6 mm (right column in each composite) posterior to the bregma to generate images containing the neocortex/hippocampus and cerebellum/brainstem, respectively. PET images are superimposed on individual MRI data, and voxel values represent the SUVR generated using the cerebellum as reference regions for each radiotracer. (**g, h**) Time-radioactivity curves (**g**; SUV) and ratio of the radioactivity uptake to the cerebellum (**h**; SUVR) in the hippocampus of 8-9-month-old Tg (red circles) and nTg (blue circles) mice over 90 and 60 min after intravenous injection of ^18^F-PM-PBB3 (left) and ^11^C-PBB3 (right). Data are mean ± SD in three Tg or nTg animals, and the same individuals were used for a head-to-head comparison of the two radiotracers.

To assess the time course of *in-vivo* labeling of intraneuronal tau aggregates with PM-PBB3, we conducted intravital two-photon laser fluorescence microscopy with a cranial window to the somatosensory cortex of the Tg and nTg mice. Comparison of PM-PBB3 and PBB3 signals in the same field of view indicated rapid entry of these probes into the brain after intravenous probe administration, reaching tau aggregates within 5 min (Figure 2d). Quantification of the background-corrected fluorescence intensity revealed that PM-PBB3 yielded 3-fold higher peak fluorescence signals in the same neurons burdened with tau aggregate than PBB3 (Figure 2e). In contrast, no noticeable increases in fluorescence signals were produced by intravenously injected PM-PBB3 in neurons of nTg mice (Figure 2d).

The *in-vivo* performance of ^18^F-PM-PBB3 and ^11^C-PBB3 as a PET probe was then examined by a head-to-head comparison in the same mice (Figure 2f-h). ^18^F-PM-PBB3 rapidly entered the brain after intravenous administration, and the peak radioactivity uptake was 1.4-fold higher than that of ^11^C-PBB3 (Figure 2g). This was followed by a prompt washout of radioactivity from the brains of nTg mice, whereas the clearance was retarded in the Tg forebrain, reflecting radioligand binding to tau deposits. ^18^F-PM-PBB3 generated a more than 2-fold higher contrast for tau lesions in the Tg hippocampus relative to nTg controls than ^11^C-PBB3 (Figure 2h).

The high brain uptake and tau contrast by ^18^F-PM-PBB3 versus ^11^C-PBB3 were primarily attributable to its stability against bio-metabolism, since unmetabolized ^18^F-PM-PBB3 accounted for 79.9% and 97.5% of the total radioactivity in plasma and brain, respectively, in contrast to unchanged ^11^C-PBB3 accounting for 2.5% and 72.4% of the total radioactivity in plasma and brain, respectively, at 5 min after intravenous injection (Table S1). ^18^F-PM-PBB3 has been confirmed to be decomposed to a hydrophilic radiometabolite in human plasma at a slower rate than metabolizing ^11^C-PBB3 (Figure S2) (Maruyama et al., 2013).

### High-contrast PET imaging of AD and PSP tau pathologies in humans enabled by ^18^F-PM-PBB3

Encouraged by nonclinical results, ^18^F-PM-PBB3 was applied to PET imaging in human subjects. As depicted in Figure 3a, the retention of ^18^F-PM-PBB3 clearly visualized the neocortical and limbic dominance of six tau isoform accumulations in an AD patient and subcortical dominance of four-repeat tau depositions in a PSP patient with Richardson’s syndrome (PSP-Richardson), a PSP subcategory with a typical clinical phenotype (Hoglinger et al., 2017), in sharp contrast to the low radiosignals sustained in the parenchyma of elderly healthy control (HC) brains. In fact, intensification of PET signals in the parieto-temporal and posterior cingulate cortices of the AD brain and the subthalamic nucleus, midbrain, and globus pallidus of the PSP brain was in agreement with the known distribution of tau pathologies in these diseases (Figure 3a).

**Figure 3.**
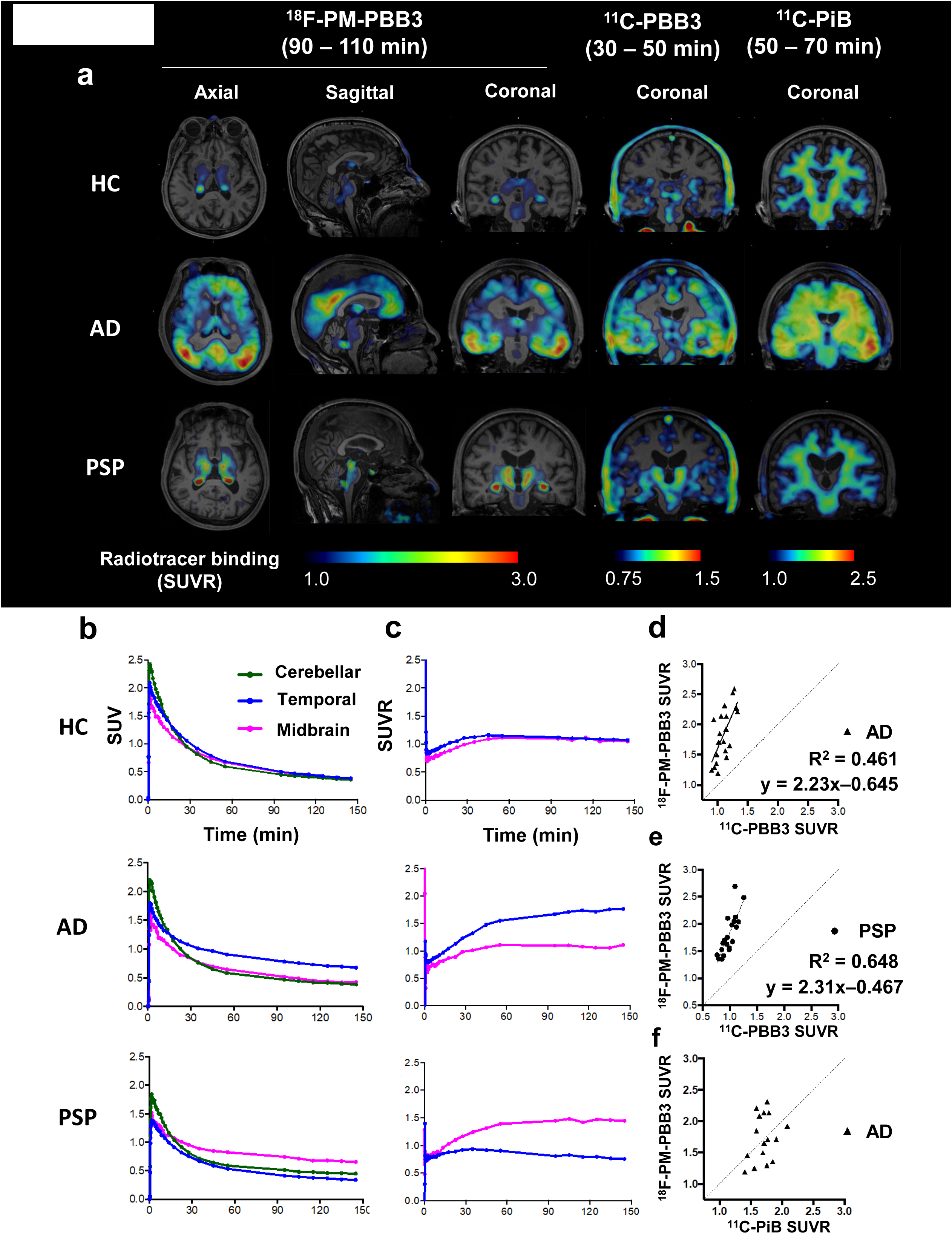
AD and PSP tau topologies visualized with high contrast by PET with ^18^F-PM-PBB3 as compared to ^11^C-PBB3 in the same human subjects. **(a)** Orthogonal ^18^F-PM-PBB3 and coronal ^11^C-PBB3 and ^11^C-PiB-PET images in the same HC, and AD and PSP patients. Images of the PSP patient were derived from an autopsy-confirmed PSP case. Data are displayed as parametric maps for radioligand SUVR. Non-thresholded ^18^F-PM-PBB3 images are also shown in Supplemental Figureure S8. (**b, c**) Time-course changes of the radioligand uptake (**b**; %SUV) and SUVR to the cerebellum (**c**) in the cerebellum (green), temporal cortex (blue) and midbrain (magenta) of representative HC, and AD and PSP patients over 150 min after intravenous injection of ^18^F-PM-PBB3. (**d-f**) Scatterplots demonstrating head-to-head comparisons of SUVR values between ^18^F-PM-PBB3 and ^11^C-PBB3 in the AD (**d**) and PSP (**e**) brains, and between ^18^F-PM-PBB3 and ^11^C-PiB in the AD brain (**f**). Significant regression results are also shown. SUVR values were generated from four VOIs in each patient.

The uptake of ^18^F-PM-PBB3 peaked rapidly after radioligand injection and subsequently declined by more than 50% across all regions of the HC brain in the next 30 min, resulting in uniformly low radioligand retention (Figure 3b). The clearance of ^18^F-PM-PBB3 was profoundly slowed in tau-burdened areas of the AD and PSP brains, conceivably reflecting the specific radioligand binding to tau aggregates (Figure 3b). The cerebellum was included in brain areas with the lowest radioactivity retention (Figure 3b), supporting the use of cerebellar gray matter as a reference tissue with a minimal tau fibril load for quantification of the radioligand binding. The target-to-reference ratio of the radioactivity (standardized uptake value ratio; SUVR) was progressively increased in affected brain areas until ~60 min after radioligand injection, and then it almost plateaued at ~90 min (Figure 3c).

To examine the superiority of ^18^F-PM-PBB3 to ^11^C-PBB3 as a high-sensitivity tau PET probe, we carried out a head-to-head comparison of PET data with these radioligands in the same individuals. The peak uptake of ^18^F-PM-PBB3 in the brain (Figure 3b) was approximately 2-fold higher than that of ^11^C-PBB3 (Figure S3), and nonspecific radioactivity retentions in the basal ganglia and venous sinuses at high levels and several neocortical areas at low levels were provoked in a HC subject by ^11^C-PBB3 but not ^18^F-PM-PBB3 (Figure 3a). Meanwhile, radioactivity accumulations in the choroid plexus, which were documented in the use of other tau radioligands including ^18^F-flortaucipir (Ikonomovic et al., 2016; Lee et al., 2018; Lowe et al., 2016), were augmented in ^18^F-PM-PBB3-PET images as compared to PET data with ^11^C-PBB3 (Figure 3a). There was a significant correlation between regional SUVRs for ^18^F-PM-PBB3 and ^11^C-PBB3 in the cortical volumes of interest (VOIs) in the AD brain (*r* = 0.679, *p* = 0.001) and subcortical VOIs in the PSP brain (*r* = 0.805, *p* < 0.001) (Figure 3d, e; see Figure S4 for details of the VOI definition), and the linear regression slopes indicated that ^18^F-PM-PBB3 produced more than 2-fold higher contrasts for AD and PSP tau deposits than ^11^C-PBB3 (Figure 3d, e).

As there was no significant correlation between SUVRs for ^18^F-PM-PBB3 and ^11^C-PiB in AD patients (*r* = 0.295, *p* = 0.268) (Figure 3f), it is unlikely that PET data with ^18^F-PM-PBB3 can be considerably affected by its cross-reactivity with Aβ deposits.

### The utility of ^18^F-PM-PBB3 for PET assessments of the topology and stage of AD-spectrum and PSP tau pathologies

We performed tau and Aβ PET scans with ^18^F-PM-PBB3 and ^11^C-PiB, respectively, for three mild cognitive impairment (MCI) and 14 AD patients (mean age ± SD, 70.7 ± 11.9 years) as well as 23 HCs (mean age ± SD, 65.2 ± 7.9 years) in order to investigate the ability of ^18^F-PM-PBB3 to capture the advancement of AD-spectrum tau pathologies in each individual. To this aim, we defined composite VOIs according to Braak’s NFT stages (Figure S4) (Cho et al., 2016; Scholl et al., 2016). The tau pathologies indicated by individual PET data were classified into stages zero (unaffected stage; 22 HCs), I/II (transentorhinal stage; one HC), III/IV (limbic stage; one MCI and three AD patient), and V/VI (neocortical stage; two MCI and 11 AD patients) by identifying Braak’s stage composite VOIs with a regional Z score > 2.5. All HCs were negative for ^11^C-PiB-PET, and all MCI and AD patients were positive for ^11^C-PiB-PET, judging from visual inspection of the acquired images. Representative ^18^F-PM-PBB3-PET images demonstrated expansions of radiosignals from the medial temporal cortex to the other neocortical and limbic areas, along with progression of the NFT stage (Figure 4a, b). ^18^F-PM-PBB3 SUVRs in stage I/II VOI were elevated in a subset of HCs, being overlapped with the values in MCI and AD patients, and this may imply accumulations of tau fibrils in the medial temporal cortex at a preclinical stage of AD or PART (Figure 4c). By contrast, SUVRs in stage III/IV and V/VI VOIs were much less variable among HCs, and all 17 AD-spectrum (MCI + AD) cases exhibited increased SUVR beyond the HC range in either of these VOIs (Figure 4c). Moreover, the radioligand accumulation in stage V/VI VOI (Figure 4c) was significantly correlated with the severity of dementia as assessed by Clinical Dementia Rating Sum of Boxes (CDRSoB) (*r* = 0.671, *p* = 0.003) (Figure 4d), whereas other stage VOIs did not show significant correlations. These results indicate that ^18^F-PM-PBB3 could detect tau depositions at preclinical and prodromal stages in the AD spectrum and PART and that the formation of ^18^F-PM-PBB3-positive tau fibrils is intimately associated with functional deteriorations of neocortical neurons in subjects with cognitive declines.

**Figure 4.**
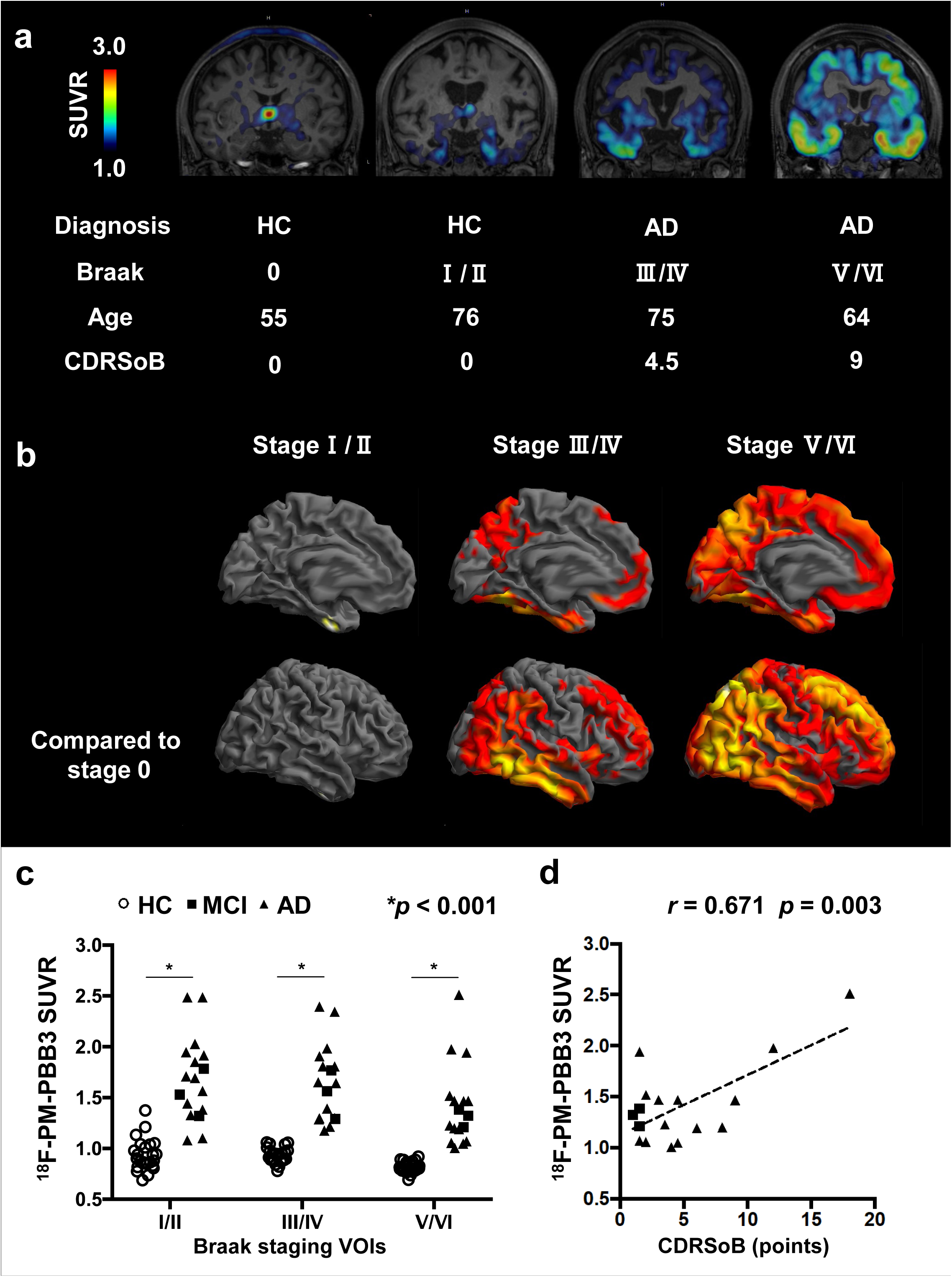
Associations between the clinical disease severity and the extent of areas showing increased ^18^F-PM-PBB3 binding in the AD spectrum. **(a)** Coronal ^18^F-PM-PBB3-PET images of HCs and AD patients classified into different Braak tau stages. **(b)** The topology of increased ^18^F-PM-PBB3 binding in subjects at each Braak stage compared to 22 HCs (stage zero). *p* <0.005, uncorrected, for one HC (stage I/II); *p* <0.05, family-wise error corrected at cluster level, for four MCI/AD patients (stage III/IV) and for 13 MCI/AD patients (stage V/VI). **(c)** Comparisons of ^18^F-PM-PBB3 binding in Braak stage VOIs between 23 HCs (white circles) and three MCI (black squares) and 14 AD (black triangles) cases. *, *p* <0.001 by two-sample t test. **(d)** Correlation of ^18^F-PM-PBB3 binding in the Braak stage V/VI VOI with CDRSoB points in MCI (black squares) and AD (black triangles) patients. *r =* 0.671 and *p =* 0.003 by Pearson’s correlation analysis. Associations between the clinical disease severity and the extension of ^18^F-PM-PBB3 binding among MCI/AD patients.

The utility of ^18^F-PM-PBB3 for evaluations of four-repeat tau pathologies was also examined in 16 PSP-Richardson patients who were negative for ^11^C-PiB-PET (mean age ± SD, 71.5 ± 6.5 years). Severities of the disease in these cases were assessed using PSP Rating Scale (PSPRS), which is a sensitive measure to evaluate global disability, including the activity of daily living, motor, and mental disabilities, and to predict prognosis in clinical practice (Golbe and Ohman-Strickland, 2007). High ^18^F-PM-PBB3 retention was observed in the subthalamic nucleus and midbrain of PSP patients relative to HCs, was progressively intensified within these subcortical structures, and was expanded to the neocortical area, including the gray and white matter of the primary motor cortex together with increase in PSPRS scores (Figure 5a). Voxel-wise PET and magnetic resonance imaging (MRI) assays revealed high spatial accordance of the radioligand accumulation and brain atrophy in the subthalamic nucleus and midbrain as compared to HCs (Figure 5b). VOI-based analyses also showed significant elevations of the radioligand SUVRs in the subcortical areas of PSP patients compared to HCs (Figure 5c). In particular, SUVR in the subthalamic nucleus, which is one of the regions most severely affected by PSP tau pathologies (Williams et al., 2007), was increased in all PSP cases with little overlap with HC values (Figure 5c), and was closely significantly correlated with PSPRS scores (Figure 5d) (*r* = 0.566, *p* = 0.018).

**Figure 5.**
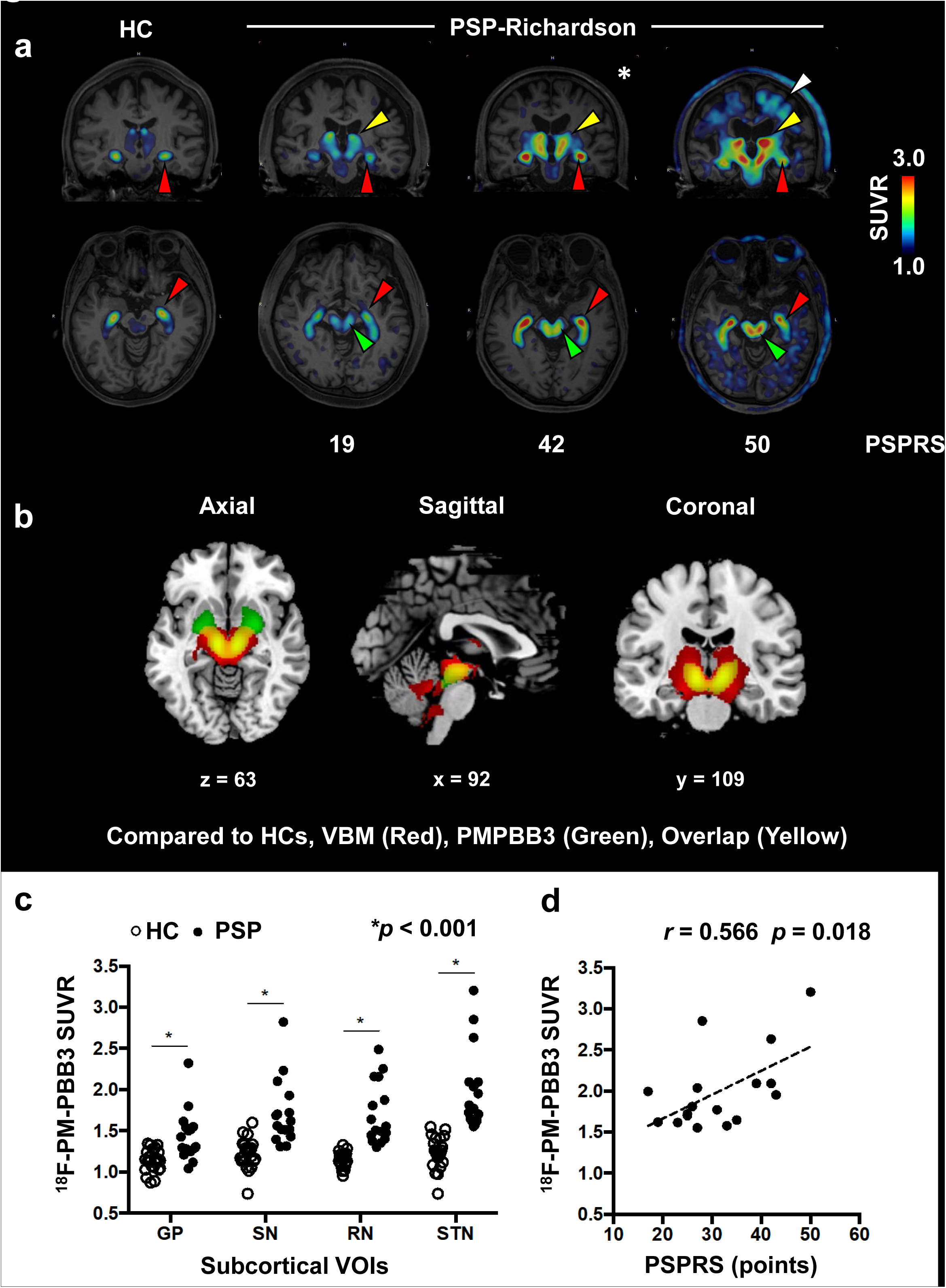
Associations between clinical disease severity and the extension of ^18^F-PMPBB3 binding in PSP-Richardson patients. **(a)** Coronal (upper) and axial (lower) ^18^F-PM-PBB3-PET images of HC and PSP21 Richardson patients with different disease severities scored by PSPRS. The red arrowheads point to the choroid plexus. PSP patients showed intensification of ^18^F-PM23 PBB3 binding in the subthalamic nucleus and neighboring thalamic and basal ganglia areas (yellow arrowhead) and midbrain (green arrowhead) and expansion to the primary motor and adjacent cerebral cortices containing white matter (white arrowhead) along with the clinical advancement. The asterisked image was derived from an autopsy confirmed PSP case. **(b)** Voxel-based analyses of brain atrophy (voxel-based morphometry, VBM; red), ^18^F-PM-PBB3 signal increase (green), and their spatial overlaps (yellow) in PSP-Richardson patients relative to HCs (*p* <0.05, family-wise error corrected at cluster level). Statistical maps are displayed in the Montreal Neurological Institute coordinate space. **(c)** Comparisons of ^18^F-PM-PBB3 uptake in subcortical VOIs, including the globus pallidus (GP), substantia nigra (SN), raphe nucleus (RN), and subthalamic nucleus (STN) between 23 HCs (white circles) and 16 PSP-Richardson patients (black circles). *, *p* <0.001 by two-sample t test. **(d)** Correlation of ^18^F-PM-PBB3 SUVR values in the STN with PSPRS points. *r* = 0.566 and *p* = 0.018 by Pearson’s correlation analysis.

### Intraindividual links between ^18^F-PM-PBB3 PET data and tau pathologies in biopsy and autopsy brain tissues proven in CBD, PSP and PiD patients

We obtained histopathological evidence that *in-vivo* ^18^F-PM-PBB3 binding reflects the abundance of three- and four-repeat tau inclusions in patients with biopsy- and autopsy-confirmed FTLD tauopathies. A clinical phenotype case of corticobasal syndrome (CBS) underwent a brain biopsy to investigate the presence of a tumor in consideration of a low-intensity lesion discovered by T1-weighted MRI as reported by Arakawa *et al*. (Arakawa et al., 2020) (Figure S5). Neuropathological and biochemical examinations of the biopsied sample from the middle frontal gyrus revealed the presence of anti-four-repeat tau-specific antibody (RD4) and/or GB positive astrocytic plaques, ballooned neurons, neuropil threads, and coiled bodies resulting from the formation of insoluble four-repeat tau aggregates, which are characteristic of CBD (Arakawa et al., 2020) (Figure 6a). Subsequent PET scans of this case showed negativity for ^11^C-PiB and notable increases of ^18^F-PM-PBB3 retentions in the primary motor cortex, basal ganglia, and brainstem consistent with the regional localization of CBD tau pathologies (Kouri et al., 2011), and middle frontal gyrus (Figure 6a and S5)

**Figure 6.**
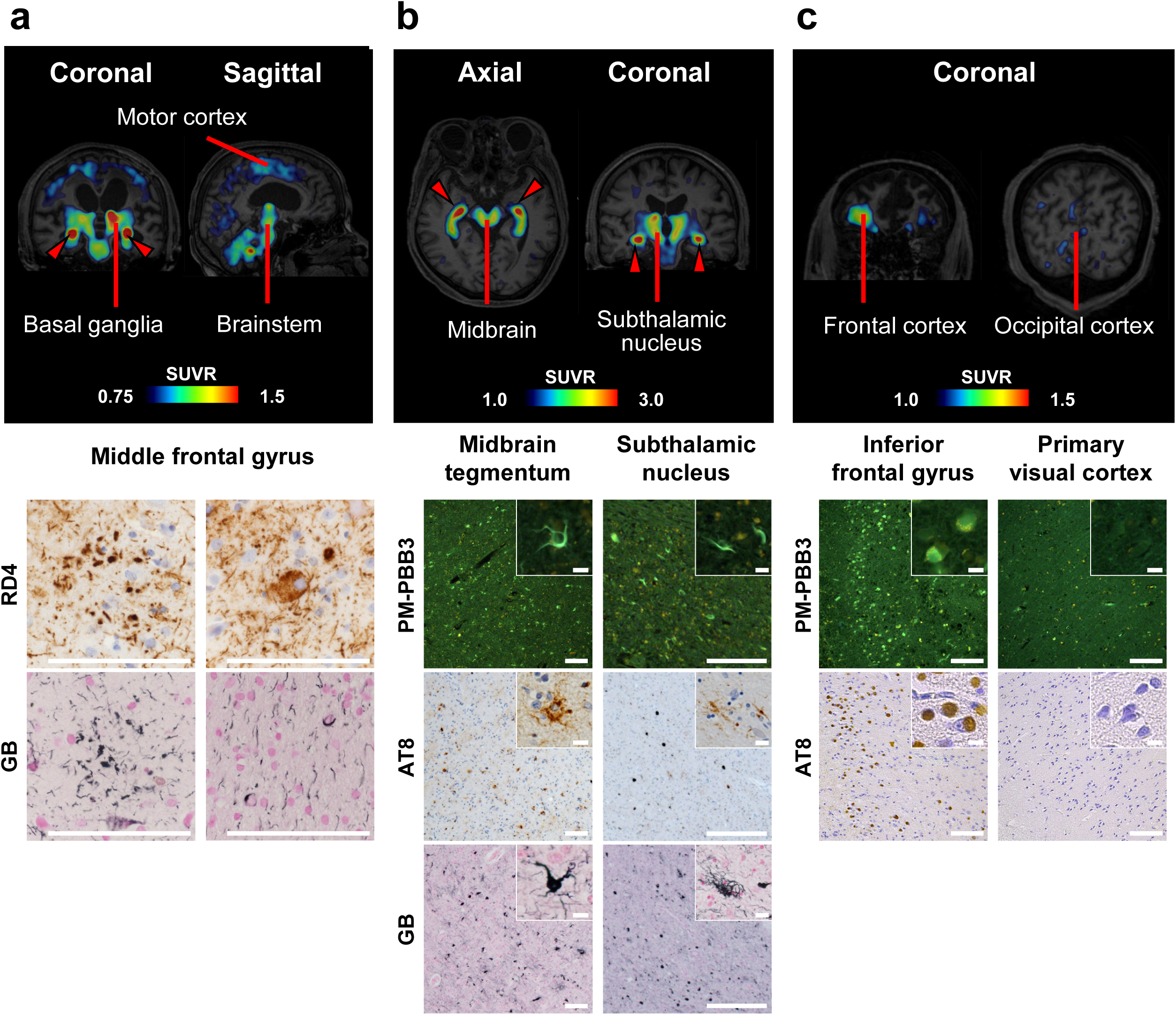
PET images of ^18^F-PM-PBB3 retentions in patients with biopsy-confirmed CBD and autopsy-confirmed PSP and PiD. (**a**) Coronal and sagittal brain images of a 68-year-old subject clinically diagnosed as having CBS (upper panels). Enhanced radioligand binding was observed in the primary motor and adjacent cortices and subcortical regions, including basal ganglia, subthalamic nucleus, midbrain, pons and choroid plexus (red arrowheads). Neuropathological assays of biopsied tissues collected from the middle frontal gyrus revealed the existence of astrocytic plaques, ballooned neurons and coiled bodies stained with RD4 and/or GB in the cortex and the corticomedullary junction (lower panels), in agreement with CBD tau pathologies. (**b**) Axial and coronal ^18^F-PM-PBB3 PET images of a 65-year-old patient with a clinical diagnosis of PSP-Richardson (upper panels). The radioligand binding was augmented in the midbrain, subthalamic nucleus, neighboring subcortical structures and choroid plexus (red arrowheads). Brain autopsy conducted two years after the PET scan demonstrated abundant accumulation of tufted astrocytes stained with non-radiolabeled PM-PBB3, AT8, and GB in the midbrain tegmentum and subthalamic nucleus (lower panels), indicating PSP as a definite diagnosis of this individual. (**c**) Coronal ^18^F-PM-PBB3 PET images of a 59-year-old patient clinically diagnosed with bvFTD (upper panels). Accumulations of radiosignals were noticeable in the frontal cortex, in contrast with the lack of radioligand binding in the occipital cortex. Brain autopsy was carried out one year after the PET scan, showing great abundance of Pick bodies and neuropil threads stained with non-radiolabeled PM-PBB3 and AT8 in the inferior frontal gyrus (lower panels). This was in sharp distinction from the few tau pathologies in the primary visual cortex (lower panels), collectively supporting a definite diagnosis of this case as PiD. Scale bars, 10 μm (inset), and 100 μm.

Brain autopsy was also performed for a PSP-Richardson patient who had received an ^18^F-PM-PBB3 PET scan (see Supplemental Materials for clinical information), and a definitive diagnosis of PSP (Cairns et al., 2007) was made on the basis of neuropathological observations. *In-vivo* ^18^F-PM-PBB3 radiosignals in the brain parenchyma were primarily concentrated in the subthalamic nucleus and midbrain (Figures 3a, 5a (asterisked images), 6b and 7 (top row)). Histochemical and immunohistochemical analyses of the autopsied specimen identified a high abundance of GB- and AT8-stained tufted astrocytes in the tegmentum and substantia nigra of the midbrain and subthalamic nucleus, and these tau inclusions were fluorescently labeled with nonradioactive PM-PBB3 (Figures 6b). In addition, GB-positive tufted astrocytes were also distributed with high abundance in the globus pallidus and lower abundance in the cerebral crus, internal capsule, and thalamus, which could contribute to the subcortical PET signals (Figures S6).

**Figure 7.**
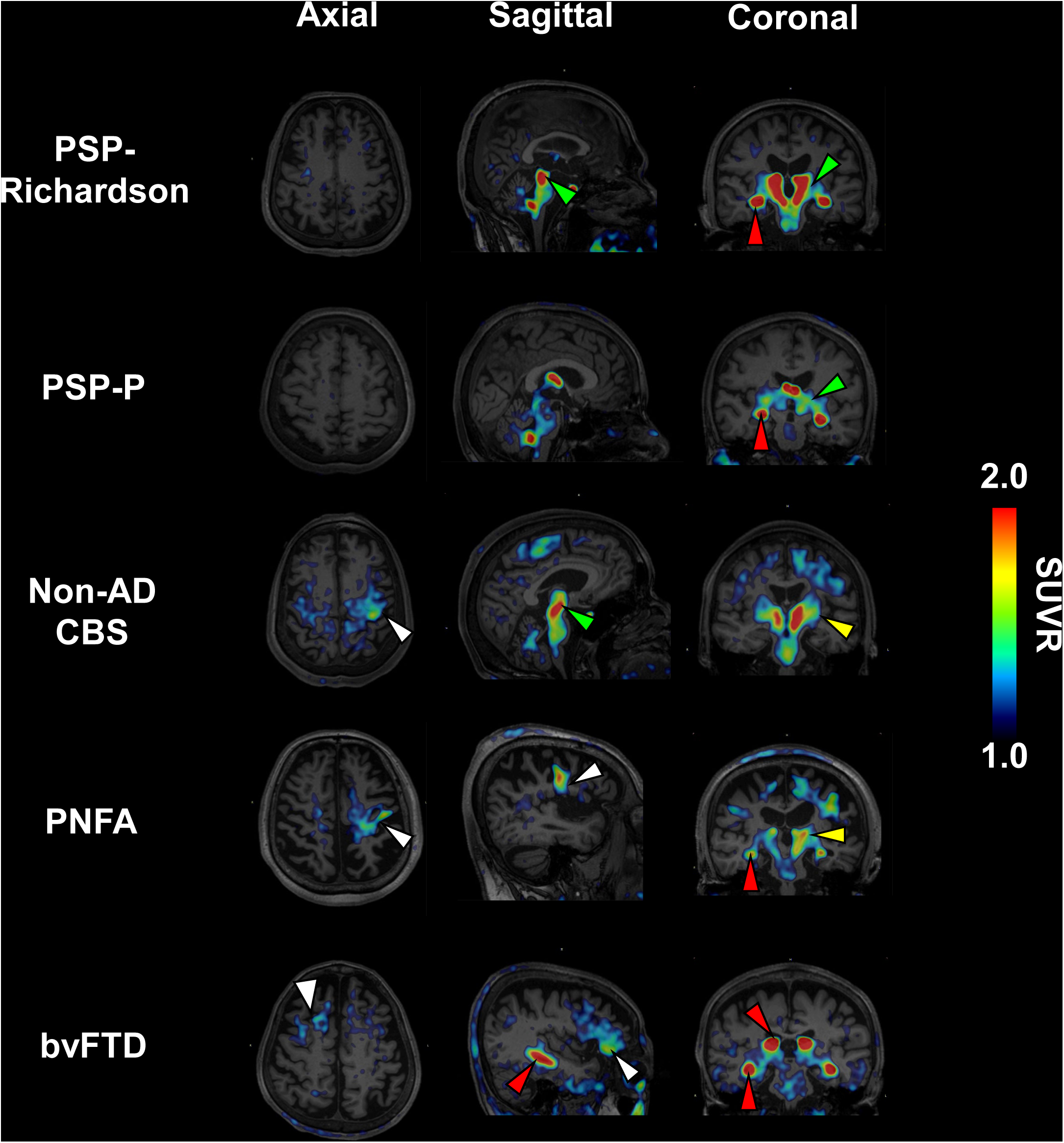
The topology of *in-vivo* ^18^F-PM-PBB3 binding in patients with diverse clinical subtypes of FTLD. Areas with intensified radiosignals, including the primary motor and cortices (white arrowheads), basal ganglia (yellow arrowheads), subthalamic nucleus/midbrain (green arrowheads) and choroid plexus (red arrowheads), are indicated in orthogonal ^18^F-PM-PBB3 PET images of individual patients. From top to bottom: a 65-year-old male clinically diagnosed with PSP-Richardson and a PSPRS score of 42 points, and also autopsy-confirmed PSP; a 62-year-old female clinically diagnosed with PSP-P and a PSPRS score of 20 points; a 65-year-old female clinically diagnosed with non-AD CBS and an MMSE score of 27 points; a 75-year-old male clinically diagnosed with PNFA and an MMSE score of 30 points; and a 72-year-old female clinically diagnosed with bvFTD and an MMSE score of 11 points.

Moreover, we conducted brain autopsy of a patient with the clinical diagnosis of behavioral variant frontotemporal dementia (bvFTD), who had undergone an ^18^F-PM-PBB3 PET scan (see Supplemental Materials for clinical information). Neuropathological examinations of the autopsied tissues provided a definitive diagnosis of PiD (Cairns et al., 2007). Increased retentions of ^18^F-PM-PBB3 were noticeable in frontal and temporal cortices, but not in the occipital cortex (Figure 6c). Histopathological assays showed numerous intraneuronal Pick bodies along with neuropil threads were doubly labeled with AT8, and PM-PBB3 fluorescence in the inferior frontal gyrus, in contrast to noticeable pathologies in the primary visual cortex (Figure 6c). Taken together, these data on imaging-pathology relationships within a subject strongly support the capability of ^18^F-PM-PBB3 for high-contrast visualization of three- and four-repeat tau deposits in the FTLD spectrum.

### Individual-based assessments of tau pathologies in living patients with diverse FTLD phenotypes

To test the feasibility of ^18^F-PM-PBB3 for evaluations of FTLD tau pathologies on an individual basis, patients with diverse clinical FTLD phenotypes (see Supplemental Materials for clinical information) were scanned with this radioligand (Figure 7). The absence of overt AD pathologies was confirmed by the negativity for ^11^C-PiB-PET in all these cases. As compared to a case included in the above-mentioned PSP-Richardson group (top row in Figure 7), a patient with PSP parkinsonism (PSP-P), which is clinically characterized by mild motor disability relative to PSP-Richardson, showed a modestly increased tracer uptake confined to the subthalamic nucleus (second row in Figure 7). A patient clinically diagnosed as non-AD CBS showed high radioligand binding in the primary motor cortex including below white matter and subcortical regions, such as the subthalamic nucleus, globus pallidus, and midbrain, with left-right asymmetry, which was predominant on the side contralateral to the more affected body side (third row in Figure 7). These changes indicate the existence of CBD tau pathologies underlying the symptomatic manifestation of CBS. Similarly, left-side dominant enhancements of radioligand retention were observed in gray and white matter of the primary motor cortex (i. e. precentral gyrus), and subcortical structures of a patient with progressive non-fluent aphasia (PNFA) (fourth row in Figure 7). It is accordingly probable that verbal symptoms in this individual represented by anarthria were chiefly attributable to CBD tau pathologies involving an inferior portion of the left precentral gyrus. Furthermore, a patient with bvFTD presented elevations of ^18^F-PM-PBB3 uptake in the lateral superior frontal gyrus and prefrontal cortex with little involvement of the primary motor cortex and subcortical structures (bottom row in Figure 7). Taken together, the tracer topographies matched with neuroanatomical variabilities of the FTLD spectrum, indicating that ^18^F-PM-PBB3 could provide an accurate diagnosis based on the evaluation of pathological backgrounds on an individual basis.

In light of the current PET observations, we constructed a schematic map illustrating that the topology of ^18^F-PM-PBB3 radiosignals is indicative of PSP, CBD, and PiD pathologies as bases of five different clinical phenotypes of FTLD (Figure 8; clinicopathological relationships were modified from Williams and Lees, 2009 (Williams and Lees, 2009)). The subcortical dominance of tau depositions characterizes pathological changes in PSP, whereas a spatial expansion of areas with ^18^F-PM-PBB3-positive tau lesions to neocortical gray and white matter centralized at the primary motor cortex may occur with disease progression. More widespread and intense accumulations in the neocortex, often with left-right asymmetry, could suggest the presence of CBD pathologies provoking clinical manifestations of CBS and PNFA. CBD tau abnormalities may also give rise to bvFTD phenotypes but enhanced PET signals in the frontal and temporal cortices with fewer involvements of the primary motor cortex and subcortical regions imply bvFTD due to PiD pathologies. Hence, PET imaging with ^18^F-PM-PBB3 potentially offers identification of the tau neuropathology linked to the clinical features of FTLD in each individual case.

**Figure 8.**
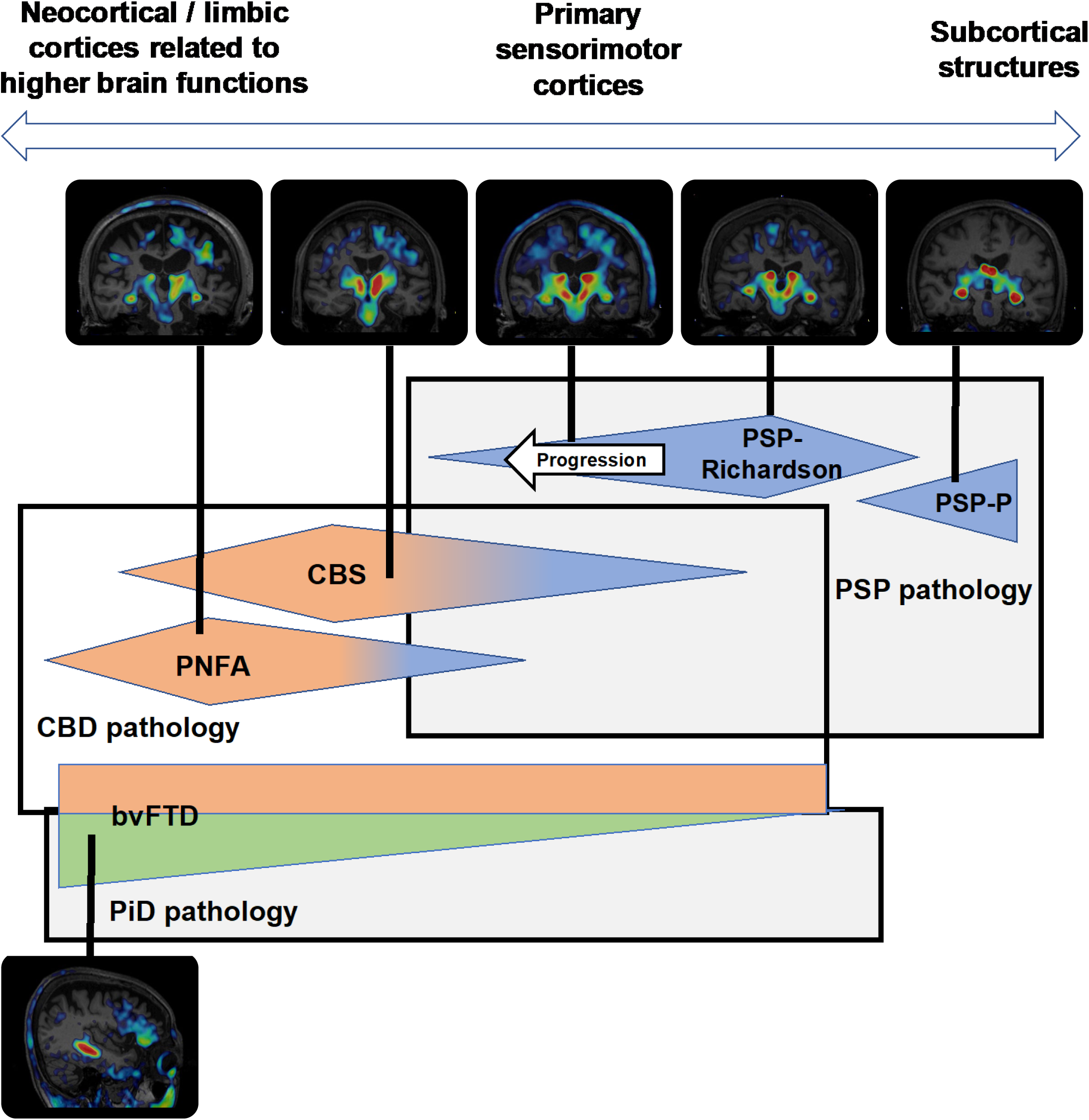
A schematic presentation of PET-detectable tau topologies in association with clinical and neuropathological nosologies of FTLD syndromes. Three tau neuropathologies underlie five clinical phenotypes, and the neocortex-to-subcortex gradient of tau depositions varies as a function of clinicopathological entity and progression of the disease. Patients whose symptomatic manifestations are confined to parkinsonism are likely to exhibit ^18^F-PM-PBB3 binding localized to subcortical areas (rightward), while patients with cortical symptoms such as apraxia and aphasia may frequently display the radioligand binding primarily in the frontotemporal cortex (leftward).

## DISCUSSION

Diagnostic evaluations of neurodegenerative tauopathies have been impeded by the lack of one-to-one associations between diverse neuropathological and clinical phenotypes. PET imaging with our novel radioligand, ^18^F-PM-PBB3, has been proven to capture a wide range of tau fibrils with different isoform compositions, conformations, and ultrastructural dimensions with contrast and dynamic range adequate for individual-based assessments of AD- and FLTD-spectrum syndromes. Of note is that imaging-neuropathology relationships within single subjects undergoing biopsy or autopsy provided compelling evidence for the ability of the present PET technology to detect tau deposits in CBD, PSP, and PiD. In addition, sensitive detection of tau inclusions in a tauopathy model mouse was enabled with a cellular scale by intravital two-photon laser microscopy and non-labeled PM-PBB3 and with a regional scale by PET and ^18^F-PM-PBB3. This multi-scale imaging system could prove useful for non-clinical investigations of neuropathologies, which can be combined with functional analyses exemplified by microscopic calcium assays and macroscopic functional MRI to clarify links between tau accumulations and neuronal dysfunctions from single-cell to brain-wide levels.

PBB derivatives exhibit unique features represented by high reactivity with three-repeat or four-repeat tau assemblies in FTLD patients and mouse models (Maruyama et al., 2013; Ono et al., 2017), in contrast to weak *in-vitro* and *in-vivo* labeling of these aggregates with flortaucipir and its ‘second-generation’ analogs (Aguero et al., 2019; Hostetler et al., 2016; Leuzy et al., 2020; Ono et al., 2017; Schonhaut et al., 2017). One of the flortaucipir derivatives, ^18^F-PI-2620, was reported to react with four- and three-repeat tau inclusions in FTLD brains (Kroth et al., 2019; Matthias Brendel, 2019), but its capability to sensitively visualize FTLD-spectrum tau pathologies has been controversial because of a lack of compelling evidence for the agreement of radioligand retentions with PSP, CBD, and PiD tau topologies. PM-PBB3 was shown to more efficiently enter the brain than the original compound, PBB3, primarily owing to its higher stability against metabolic conversions. The substantial uptake of unmetabolized PM-PBB3 in the brain resulted in the visualization of tau lesions with a high dynamic range, which was more than 2-fold of the value yielded by PBB3 in human brains. Besides the pharmacokinetic properties, low retentions of radiosignals in venous sinuses and putative penetration vessels in the striatum facilitate the identification of pathological changes in the use of ^18^F-PM-PBB3 as compared with ^11^C-PBB3 (Maruyama et al., 2013). Moreover, ^18^F-PM-PBB3 was not reactive with MAO-A and MAO-B and induced no increases in the striatal radioactivity related to MAO-B, unlike ^18^F-THK5351 and allied quinoline derivatives (Harada et al., 2017; Ng et al., 2017). This characteristic is also beneficial for tau PET imaging without off-target radioligand binding in MAO-B-expressing reactive astrocytes (Carter et al., 2012).

It is noteworthy that intensification and expansion of tau depositions in association with the progression of AD and PSP could be illustrated by ^18^F-PM-PBB3 PET, indicating that this radioligand could illuminate tau species critically involved in deteriorations of neuronal functions. Similar to previous indications of prion-like tau dissemination in AD brains capturable by PET with other probes (Betthauser et al., 2020; Cho et al., 2016; Jack et al., 2018; Leuzy et al., 2020; Pascoal et al., 2018; Shimada et al., 2017), ^18^F-PM-PBB3 was capable of visualizing the spatial spread of tau depositions in line with Braak’s tau staging, with regional radioligand retentions correlated with clinical advancements assessed by CDRSoB. Significantly, enhanced radiosignals in the subthalamic nucleus were concurrent with the symptomatic advancement of PSP scored by PSPRS and were expanded from subcortical to neocortical areas, seemingly in accordance with emergences of cognitive deficits. This observation may be attributable to the propagation of tau fibrillogenesis along with corticostriatal and corticothalamic connections, although prionoid properties of PSP-type tau will need to be proven by longitudinal PET tracking of tau aggregates in the same individuals.

The current data also provide evidence for the *in-vivo* performance of ^18^F-PM-PBB3 as a diagnostic adjunct to the identification and differentiation of various clinical FTLD subtypes on a neuropathological basis. Indeed, distinctions between AD/PART and CBD/PSP in PNFA and CBS and among AD/PART, CBD/PSP, and PiD in bvFTD were allowed for each case according to the topology of PM-PBB3-positive tau deposits. Since these clear separations have not been possible with the use of previous tau PET ligands (Endo et al., 2019; Schonhaut et al., 2017) and other imaging modalities such as volumetric MRI, the current technology paves the way for the construction of a biomonitoring system for the selection of an adequate disease-modifying therapeutic. This advantage will be of particularly great importance for the development and implementation of treatments against glial deteriorations in consideration of astrocytic and oligodendrocytic tau accumulations that predominate PSP and CBD. It is yet to be clarified whether PSP and CBD are precisely differentiated in terms of regional distribution and laterality of PET-detectable tau lesions, notwithstanding the fact that these two disease categories occasionally display considerable overlaps at clinical and neuropathological levels (Dickson, 1999).

To date, the application of cryo-electron microscopy has led to the revelation that tau filaments are constituted of a disease-specific self-assembling portion, providing intrinsic binding pockets for PET ligands (Goedert et al., 2018; Murugan et al., 2018). This diversity of the interaction between distinct fibrils and ligands may not necessarily arise from differences in the tau isoform composition but could stem from additional conformational variations. In fact, tau assemblies in chronic traumatic encephalopathy (CTE) have been found to be composed of a conformer distinct from the AD-type tau filament (Falcon et al., 2019), despite the incorporation of all six isoforms in both AD and CTE tau deposits. This could result in differential affinities of tau PET probes for the two tau fibril species, and our pilot PET study has demonstrated sensitive detection of CTE pathologies with ^18^F-PM-PBB3 (Takahata et al., paper in preparation), which has not been achieved by ^11^C-PBB3 (Takahata et al., 2019) and flortaucipir (Mantyh et al., 2020; Stern et al., 2019).

Along with technical benefits, several issues should also be considered regarding the utilization of ^18^F-PM-PBB3 in clinical PET scans. The accumulation of radioactivity in the choroid plexus might hinder quantitative assessments of tau depositions in neighboring structures, including the hippocampus, although this did not overtly influence the measurement of radioligand retentions in the parahippocampal gyrus, which is the area involved in tau pathologies at the earlies Braak stage. Radiosignals in the choroid plexus were documented in previous works with several other tau radioligands (Ikonomovic et al., 2016; Johnson et al., 2016) and are likely derived from compounds bound to Biondi bodies mainly composed of non-tau but yet unidentified proteins (Ikonomovic et al., 2016; Lowe et al., 2016). Another caveat might be photoisomerization of PM-PBB3, which is similar to the reported property of PBB3 (Hashimoto et al., 2014). Our data indicated that the conversion of PBB3 and PM-PBB3 to their isomers could be entirely blocked using a UV-free LED light in the radiosynthesis and administration to subjects, requiring small additional equipment in nuclear medicine facilities.

The translational research workflow with PM-PBB3 offers a seamless evaluation of candidate anti-tau therapeutics (Congdon and Sigurdsson, 2018; Shoeibi et al., 2018) in non-clinical and subsequent clinical settings. Comparisons between efficacies of such potential drugs in animal models and humans with the same imaging-based biomarker will also help refine these models in view of their resemblance to tauopathy cases.

To our conclusion, the new bimodal imaging agent, PM-PBB3, enabled high-contrast optical and PET detection of diverse tau conformers at cellular, regional, and global scales in animal brains. This probe also captured AD- and FTLD-type tau pathologies with a dynamic range sufficient for differentiation and staging of tauopathy subtypes in each subject, reinforcing investigations of the neuropathological basis of clinical phenotypes in living tauopathy cases.

## Data Availability

The authors confirm that the data supporting the findings of this study are available within the article and its supplementary materials.

## ACKNOWLEDGEMENTS

The authors thank all patients and their caregivers for participation in this study, Kazuko Suzuki, Shizuko Kawakami and Momoyo Takanashi for their assistance as clinical coordinators, Yasuharu Yamamoto, Yoshikazu Nakano, Takamasa Maeda and the members of the Translational Team for their support with the PET scans, Hiromi Sano and Naoto Sato for their support with the MRI scans, Jun Maeda, Bin Ji, Takeharu Minamihisamatsu, Shoko Uchida and Kana Osawa for their support with mouse studies, the staff of the Department of Radiopharmaceuticals Development for their radioligand synthesis and metabolite analysis, and Izumi Kaneko, Chieko Kurihara and Atsuo Waki for the monitoring and audit of the study. We thank John Q. Trojanowski and Virginia MY. Lee at the Center for Neurodegenerative Disease Research and Institute on Aging, Perelman School of Medicine, University of Pennsylvania, and Akiyoshi Kakita and Hiroshi Shimizu at the Department of Pathology, Brain Research Institute, Niigata University, for kindly sharing postmortem brain tissues, Masato Hasegawa at the Department of Dementia and Higher Brain Function, Metropolitan Institute of Medical Science for the biochemical analysis, and APRINOIA Therapeutics Inc. for kindly sharing precursor of ^18^F-PM-PBB3. The authors acknowledge support for the recruitment of patients by Shigeki Hirano and Satoshi Kuwabara at the Department of Neurology, Graduate School of Medicine, Chiba University; Taku Hatano, Yumiko Motoi, Shinji Saiki and Nobutaka Hattori at the Department of Neurology, Juntendo University School of Medicine; Morinobu Seki, Hajime Tabuchi and Masaru Mimura at the Department of Neuropsychiatry, Keio University School of Medicine; Ikuko Aiba at the Department of Neurology, National Hospital Organization Higashinagoya National Hospital; Yasushi Shiio and Tomonari Seki at the Department of Neurology Tokyo Teishin Hospital.

This study was supported in part by AMED under Grant Number JP18dm0207018, JP19dm0207072, JP18dk0207026, JP19dk0207049 to M. H., and by MEXT KAKENHI Grant Number JP16H05324 to M. H., JP18K07543 to H. Shimada, JP26117011 to N. S., and by JST CREST Grant Number JPMJCR1652 to M. H.

## AUTHOR CONTRIBUTIONS

K.Tagai and M.Ono conceived the experiments and wrote the paper. Y.Takado, T.S., M.Shigeta, N.S., M.H. and H.Shimada contributed to the conception and design of the study. S.K., K.Takahata, M.K., H.Shinotoh, Y.Sano. and K.M. contributed to the clinical studies through the collection and processing of patient samples, provided clinical data, and provided insight. H.A., H.Suzuki, M.Onaya, T.T., K.A., N.Arai, N.Araki and Y.Saito contributed to the brain biopsy, autopsies and their histological examinations. C.S., Y.K. and M.I. contributed to the kinetic analysis. H.T., M.T. and Y.Tomita contributed to *in vivo* two-photon fluorescence microscopy. K.K., T.K., M Okada and M.-R.Z. contributed to the radioligand synthesis and metabolite analysis. M.Shimojo helped assay validation and writing the manuscript.

## DECLARATION OF INTERESTS

H.Shimada., M.-R.Z., T.S., and M.H. hold patents on compounds related to the present report (JP 5422782/EP 12 884 742.3/CA2894994/HK1208672).

## STAR⋆METHODS

## CONTACT FOR REAGENT AND RESOURCE SHARING

Further information and requests for resources and reagents should be directed to and will be fulfilled by the Lead Contact, Makoto Higuchi

## EXPERIMENTAL MODEL AND SUBJECT DETAILS

### Mice

The parental P301L tau responder line, parental tTA activator line, and the resultant F1 rTg4510 mice and littermates were generated and maintained as previously described (Ishikawa et al., 2018; Santacruz et al., 2005). All mice studied here were maintained and handled in accordance with the National Research Council’s Guide for the Care and Use of Laboratory Animals. Protocols for the present animal experiments were approved by the Animal Ethics Committees of the National Institute of Radiological Science. All procedures involving live mice received prior approval from the Institutional Animal Care and Use Committee of the University of Florida.

### Human subjects

We included 23 HCs and 39 patients with diverse tauopathies - AD and FTLD spectrum in the present study. All HCs were without a history of neurologic and psychiatric disorders. Three MCI patients and 14 AD patients met Petersen’s criteria (Petersen et al., 1999) and NINDS-ADRDA criteria, respectively (McKhann et al., 1984). Seventeen PSP patients were clinically diagnosed according to the Movement Disorder Society new diagnostic criteria (Hoglinger et al., 2017) and classified into each clinical variant: 16 PSP-Richardson and one PSP-P. Five other FTLD spectrum; two CBS, one PNFA and two bvFTD, were also diagnosed according to established criteria (Armstrong et al., 2013; Gorno-Tempini et al., 2011; Rascovsky et al., 2011). In the present study, HCs and FTLD spectrum patients required PiB (-) to exclude preclinical and co-pathological AD, whereas MCI and AD patients needed PiB (+) by visual assessment. In addition, diagnoses of some patients were also validated according to their neuropathological examinations. One CBS patient was confirmed with CBD according to brain tissue biopsy before the PET scan (Arakawa et al., 2020); each of the PSP-Richardson and bvFTD patients was also neuropathologically diagnosed as PSP and PiD (Cairns et al., 2007) by autopsies after two years and one year after each PET scan, respectively.

Written informed consents were obtained from all subjects and/or from spouses or other close family members when subjects were cognitively impaired. This study was approved by the Radiation Drug Safety Committee and National Institutes for Quantum and Radiological Science and Technology Certified Review Board of Japan. The study was registered with UMIN Clinical Trials Registry (UMIN-CTR; number 000030248).

## METHOD DETAILS

### Compounds and Antibodies

PM-PBB3 1 -fluoro-3 -((2-((1*E*,3*E*)-4-(6-(methylamino)pyridine-3 -yl)buta-1,3 -dien-1 - yl)benzo[*d*]thiazol-6-yl)oxy)propan-2-ol (Figure. 1a) and tosylate precursor of ^18^F-PM-PBB3 protected with tert-Butyloxycarbonyl group and 2-tetrahydropyranyl group (Figure. S1) were custom-synthesized (Nard Institute). The precursor of ^18^F-PM-PBB3 was also provided by APRINOIA Therapeutics Inc. PBB3 (2-((1*E*,3*E*)-4-(6-(methylamino)pyridine-3-yl)buta-1,3-dienyl)benzo[*d*]thiazol-6-ol) (Figure.1a) and desmethyl precursor of ^11^C-PBB3 were also custom-synthesized (Nard Institute) (Maruyama et al., 2013). The reference standard for ^11^C-PiB, 6-OH-BTA-1, is commercially available (ABX), and the desmethyl precursor of ^11^C-PiB protected with methoxymethyl group, 6-MOMO-BTA-0, was custom-synthesized (KNC Laboratories). PBB5 (Maruyama et al., 2013), BTA-1, clorgiline and selegiline are commercially available (Sigma-Aldrich). A monoclonal antibody against tau phosphorylated at Ser 202 and Thr 205 (AT8, Endogen) and four-repeat tau isoform (RD4, Upstate) are commercially available.

### Postmortem brain tissues

Postmortem human brains were obtained from autopsies carried out at the Center for Neurodegenerative Disease Research of the University of Pennsylvania Perelman School of Medicine on patients with AD, PiD, PSP and CBD, and at the Department of Neurology at the Chiba-East National Hospital on patients with PSP. Tissues for homogenate binding assays were frozen, and tissues for histochemical, immunohistochemical and autoradiographic labeling were frozen or fixed in 10% neutral buffered formalin followed by embedding in paraffin blocks.

### Radiosynthesis

^11^C-PBB3 was radiosynthesized using its desmethyl precursor, as the method previously described (Maruyama et al., 2013). Radiolabeling of ^18^F-PM-PBB3 was performed as the synthetic pathway described in Figure. S1. Tosylate precursor of ^18^F-PM-PBB3 was reacted with ^18^F-fluoride in the presence of dimethyl sulfoxide, K_2_CO_3_ and and K222 at 110°C for 15 min. After cooling the reaction vessel to 90°C, hydrochloric acid was added to the mixture and maintained for 10 min to delete the protecting groups. Sodium acetate was added to the reaction vessel, and the radioactive mixture was transferred into a reservoir for high-performance liquid chromatography (HPLC) purification (Waters Atlantis prep T3 column, 10 × 150 mm; CH_3_CN/50 mM AcONH_4_ = 4/6, 5 ml/min). The fraction corresponding to ^18^F-PM-PBB3 was collected in a flask containing 25% ascorbic acid solution and Tween 80, and was evaporated to dryness under a vacuum. The residue was dissolved in 17 ml of saline (pH 7.4) to obtain ^18^F-PM-PBB3 as an injectable solution. The final formulated product was radiochemically pure (≥ 95%) as detected by analytic HPLC (Waters Atlantis prep T3 column, 4.6 × 150 mm; CH_3_CN/50 mM AcONH_4_ = 4/6, 1 ml/min). The specific activity of ^18^F-PM-PBB3 at the end of synthesis was 58-761 GBq/μmol, and ^18^F-PM-PBB3 maintained its radioactive purity exceeding 90% for over 3 hr after formulation. Radiolabelling of ^11^C-PiB was performed as previously described (Maeda et al., 2011).

PBB3 is known to undergo photo-isomerization under ordinary fluorescent light (Hashimoto et al., 2014). ^18^F-PM-PBB3 and ^11^C-PBB3 in a colorless vial were isomerized by exposure to the fluorescent light for 30 min (Figure. S7a and b, left). UV-VIS absorption spectra for PM-PBB3 and PBB3 indicated that these compounds do not absorb light with wavelength longer than 500 nm (Figure. S7c). Then, ^18^F-PM-PBB3 and ^11^C-PBB3 in a colorless vial were placed under a UV-cut light (<500 nm wavelength cutoff, ECOHiLUX HES-YF, 2200 lm, Iris Oyama Inc.) for 30 min, and both compounds were found to be stable under this condition (Figure. S7a and b, right). Based on these results, radiosyntheses of ^18^F-PM-PBB3 and ^11^C-PBB3 and all experiments with these compounds were performed under the UV-cut light to avoid photo-isomerization of these compounds.

### *In vitro* and *ex vivo* autoradiography

*In vitro* autoradiography was performed using 6-μm-thick deparaffinized sections derived from AD and 20-μm-thick fresh frozen sections post-fixed in 4% paraformaldehyde solution derived from PSP brains. For labeling with ^18^F-PM-PBB3, sections were pre-incubated in 50 mM Tris-HCl buffer, pH 7.4, containing 20% ethanol at room temperature for 30 min, and incubated in 50 mM Tris-HCl buffer, pH 7.4, containing 20% ethanol and 5 nM of ^18^F-PM-PBB3 (specific radioactivity: 58 GBq/μmol) at room temperature for 60 min. The samples were then rinsed with ice-cold Tris-HCl buffer containing 20% ethanol twice for 2 min, and dipped into ice-cold water for 10 sec. For *ex vivo* autoradiography, Tg and nTg wild type at 9.7 months of age were anesthetized with 1.5% (v/v) isoflurane and given 33.3 MBq ^18^F-PM-PBB3 (specific radioactivity: 248.4 GBq/μmol) by syringe via tail vein. The animals were killed by decapitation at 30 min after tracer administration. Brain was harvested and cut into 20-μm-thick sections on a cryostat (HM560; Thermo Fisher Scientific).

The sections labeled with ^18^F-PM-PBB3 were subsequently dried by treating with warm air, and exposed to an imaging plate (BAS-MS2025, Fuji Film). The imaging plate was scanned with a BAS-5000 system (Fuji Film) to acquire autoradiograms. Fresh frozen sections generated in the process of *ex vivo* autoradiography were post-fixed in 4% paraformaldehyde solution for the subsequent histological examination.

### Histological examination

For fluorescence labeling, deparaffinized sections and sections used for autoradiography were incubated in 50% ethanol containing 25 μM of non-radiolabeled PM-PBB3 at room temperature for 30 min. The samples were rinsed with 50% ethanol for 5 min, dipped into distilled water twice for 3 min, and mounted in non-fluorescent mounting media (VECTASHIELD, Vector Laboratories). Fluorescence images were captured using a DM4000 microscope (Leica) equipped with a custom filter cube for PBB3 (excitation band-pass at 414/46 nm and suppression low-pass with 458 nm cutoff) (Ono et al., 2017). Following microscopy, sections were autoclaved for antigen retrieval, and immunostained with AT8. Immunolabeling was then examined using DM4000. Finally, the tested samples were used for GB staining with Nuclear Fast Red (Sigma-Aldrich) counter-staining after pretreatment with 0.25% KMnO_4_ followed by 2% oxalic acid.

### *In vivo* two-photon fluorescence microscopy

Two weeks before the measurement, surgery to create cranial windows was performed. For this procedure, the animals were anesthetized with a mixture of air, oxygen, and isoflurane (3-5% for induction and 2% for surgery) via a facemask, and a cranial window (3-4 mm in diameter) was attached over the left somatosensory cortex, centered at 1.8 mm caudal and 2.5 mm lateral from the bregma, according to the ‘Seylaz-Tomita method’ (Tomita et al., 2005). A custom metal plate was affixed to the skull with a 7-mm-diameter hole centered over the cranial window.

Sulforhodamine 101 (MP Biomedicals) dissolved in saline (10 mM) was injected intraperitoneally (8 μl/g body weight) just before initiation of the imaging experiments. The awake animals were placed on a custom-made apparatus, and real-time imaging was conducted by two-photon laser-scanning microscopy (TCS-SP5 MP, Leica) with an excitation wavelength of 900 nm. Two-photon imaging was performed before and 5, 30, 60, 90 and 120 min after intravenous injection of 0.05 mg of PM-PBB3 and PBB3 dissolved in dimethyl sulfoxide: saline = 1: 1 (0.05% W/V). An emission signal was separated by a beam splitter (560/10 nm) and simultaneously detected through a bandpass filter for sulforhodamine 101 (610/75 nm) and PM-PBB3 and PBB3 (525/500 nm). A single image plane consisted of 1024 by 1024 pixels, and in-plane pixel-size was 0.25–0.45 μm depending on an instrumental zoom factor. Images were acquired at a depth of 0.2–0.4 mm from the cortical surface. In each resulting images from Tg mouse, fluorescence intensity from 10 randomly selected fluorescence-labeled pathologies were measured by ImageJ, and the average was calculated after background normalization. It should be noted that the background intensity of each image was acquired by averaging the fluorescence intensity at 10 randomly selected areas where no fluorescence-labeled pathologies were found.

### *In vivo* PET imaging in mice

PET scans were performed using a microPET Focus 220 animal scanner (Siemens Healthcare) providing 95 transaxial slices 2.0 mm (center-to-center) apart, a 19.0-cm transaxial field of view (FOV), and a 7.6-cm axial FOV. Prior to the scans, Tg and nTg mice at 8-9 months of age (n = 3 each) were anesthetized with 1.5% (v/v) isoflurane. Emission scans were carried out for 90 min (^18^F-PM-PBB3) or 60 min (^11^C-PBB3) in 3D list mode with an energy window of 350-750 keV, immediately after intravenous injection of ^18^F-PM-PBB3 (28.3 ± 10.3 MBq) or ^11^C-PBB3 (29.7 ± 9.3 MBq). All list-mode data were sorted into 3D sinograms, which were then Fourier-rebinned into 2D sinograms (frames for ^18^F-PM-PBB3: 4 × 1, 8 × 2, and 14 × 5 min, frames for ^11^C-PBB3: 10 × 1, 6 × 5, and 2 × 10 min). Average images were generated with maximum *a posteriori* reconstruction, and dynamic images were reconstructed with filtered backprojection using a 0.5-mm Hanning filter. VOIs of hippocampus and cerebellum were placed using PMOD image analysis software (PMOD Technologies Ltd) with reference to the individual MR image.

### *In vitro* binding assay

Frozen tissues derived from the frontal cortex of an AD patient, the motor cortex of a PSP patient and the forebrain of Tg and nTg mice were homogenized in 50 mM Tris-HCl buffer, pH 7.4, containing protease inhibitor cocktail (cOmplete™, EDTA-free, Roche), and stored at -80°C pending analyses. To assay radioligand binding with homologous or heterologous blockade, these homogenates (100 μg tissue) were incubated with 1 nM ^18^F-PM-PBB3 (specific radioactivity: 257.2 ± 22.2 GBq/μmol) in the presence or absence of non-radiolabeled PM-PBB3, BTA-1, clorgiline and selegiline at varying concentrations ranging from 10^−11^-10^−6^ M in Tris-HCl buffer containing 10% ethanol, pH 7.4, for 30 min at room temperature. Non-specific binding of ^18^F-PM-PBB3 was determined in the presence of 5×10^−7^ M PM-PBB3. Samples were run in quadruplicates and specific radioligand binding was determined as pmol/g tissue. Ki was determined by using non-linear regression to fit a concentration-binding plot to one-site binding models derived from the Cheng-Prusoff equation with GraphPad Prism version 5.0 (GraphPad Software), followed by F-test for model selection. Kd and Bmax were calculated from homologous competitive binding using this function:

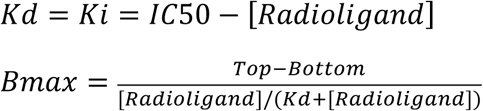

where IC50 and [Radioligand] are concentration of the competitor inducing 50% inhibition and radiotracer concentration, respectively, and Top and Bottom are upper and lower plateaus of the plot curve, respectively.

### *In vivo* MRI and PET Imaging in Human Subjects

MR images were acquired with a 3-T scanner, MAGNETOM Verio (Siemens Healthcare). Three-dimensional volumetric acquisition of a T1-weighted gradient echo sequence produced a gapless series of thin sagittal sections (TE = 1.95 ms, TR = 2300 ms, TI = 900 ms, flip angle = 9°, acquisition matrix = 256×256×250, voxel size=1×1×1mm). PET assays were conducted with a Biograph mCT flow system (Siemens Healthcare), which provides 109 sections with an axial field of view of 16.2 cm. The intrinsic spatial resolution was 5.9 mm in-plane and 5.5 mm full-width at half-maximum axially. Images were reconstructed using a filtered back projection algorithm with a Hanning filter (4.0 mm full-width at half-maximum).

^18^F-PMPBB3 had an average injected dose of 189.5 ± 22.5 MBq with a molar activity at the time of injection of 238.5 ± 71.8 GBq/μmol. ^18^F-PM-PBB3 PET scans were performed with two steps of scan protocol. Of the first protocol, dynamic PET scans with arterial blood sampling were performed with two imaging sessions of 60 min each with a 30 min break between sessions (0−60 and 90-150 min). The dynamic scan consisted of 12×10 s, 2×30 s, 7×1 min, 1×2 min, 1×3 min, 3×5 min, 3×10 min for the initial 60-min session, and 6×10-min frames for the second 60-min session. Of the second protocol, a 20-min PET acquisition was performed 90 min after injections (4×5-min frames) (see also Supplemental Materials).

^11^C-PBB3 and ^11^C-PiB PET scans were performed following a previously reported protocol (Kimura et al., 2015; Maruyama et al., 2013; Shimada et al., 2017). Seventy-minute dynamic PET scans were performed after an intravenous injection of ^11^C-PBB3 (injected dose: 423.1 ± 57.2 MBq, molar activity: 70.0 ± 18.6 GBq/ μmol). ^11^C-PiB (injected dose: 521.2 ± 87.3 MBq, molar activity: 81.8 ± 40.5 GBq/ μmol) PET scan was conducted with a 20-min acquisition 50 min after injections; a ECAT EXACT HR+ scanner (CTI PET Systems, Inc.) was also utilized for ^11^C-PiB alternatively.

### Image analyses in Human Subjects

Data preprocessing was performed using PMOD 3.8 and Statistical Parametric Mapping software (SPM12, Wellcome Department of Cognitive Neurology). Acquired PET images were rigidly coregistered to individual T1-weighted MR images. SUVR images were generated from averaged PET images at the following intervals: 30-50 min (^11^C-PBB3), 50-70 min (^11^C-PiB) and 90-110 min (^18^F-PM-PBB3) post injection, respectively. Cerebellar gray matter was used as reference region. Regarding VOI analyses, surface-based cortical reconstruction was conducted with FreeSurfer 6.0 (http://surfer.nmr.mgh.harvard.edu/) from the Desikan–Killiany–Tourville atlas (Klein and Tourville, 2012), and then cortical and Braak-staging VOIs were generated. Subcortical VOIs were transformed from a template atlas (Talairach Daemon atlas from the Wake Forest University PickAtlas version 3.0.5) to each native space using the deformation field obtained from the tissue-class segmentation of SPM12. Regarding voxel-wise analysis, each image was spatially normalized to MNI (Montreal Neurologic Institute) space using Diffeomorphic Anatomical Registration Through Exponentiated Lie Algebra (DARTEL) algorithm. Subsequently, normalized images were smoothed with a Gaussian kernel with an 8-mm full-width at half-maximum. Partial volume correction was not performed in the present study.

### Characteristics of ^18^F-PM-PBB3 in Human Subjects

We explored uptake into the brain, and the dynamic range and distribution of specific binding of ^18^F-PM-PBB3. A head-to-head comparison of ^18^F-PM-PBB3, ^11^C-PBB3 and ^11^C-PiB was conducted in the same individuals. Regional time-activity curves as standardized uptake value (SUV) and SUVR were generated over the time course of the dynamic scan; in addition, linear regression analyses were performed among the regional SUVR of each tracer derived from the same subjects. For AD, VOIs were set in each lobe of the cerebral cortex to compare the dynamic range and distribution of specific binding among the three tracers. For PSP, the same number of VOIs as for AD were set to compare the dynamic range among PBB3 compounds in the subcortical structures where PSP is considered to show moderate to high tau burden (Hauw et al., 1994) - globus pallidus, substantia nigra, red nucleus, and subthalamic nucleus. Besides, the midbrain was also used as a broad target-region for PSP (Figure. S4).

### Assessing Tau Deposits Associated with AD

Progression of tau deposits in HCs, MCI and AD patients were evaluated according to the image-based tau stage. We calculated SUVRs and Z scores of composite VOIs based on Braak’s pathological stages of neurofibrillary tangle: stages I / II (transentorhinal), III/IV (limbic) and V/VI (neocortical) (Cho et al., 2016; Scholl et al., 2016). Hippocampus was excluded from the analysis because of contamination of the signal from off-target binding in the choroid plexus. The stage showing highest regional Z score > 2.5 was assigned to the individual image-based tau stage: subjects showing lack of involvement of stages I / II were classified as stages zero. Subsequently, we assessed distribution of tau deposits and clinical association in each image-based tau stage. Voxel-level comparisons were performed comparing stages I /II, III/IV and V/VI to stage zero. Group comparisons between MCI/AD patients and HCs were also performed in each VOI; in addition, regression analyses between SUVR of each VOI and CDRSoB were also performed in MCI and AD patients.

### Assessing Tau Deposits Associated with PSP-Richardson

Associations among tau deposits, clinical symptoms and brain atrophy were assessed. Brain atrophy was estimated by voxel-based morphometry. Voxel-level comparisons between PSP-Richardson patients and HCs were performed regarding distributions of tau deposits and brain atrophy. Group comparisons using subcortical VOIs were also conducted; in addition, regression analyses between SUVR of each VOI and PSPRS scores were also performed in PSP-Richardson patients.

## QUANTIFICATION AND STATISTICAL ANALYSIS

Statistical calculations with respect to VOI analyses were performed using GraphPad Prism 7.0. Group comparisons of SUVR values derived from VOIs between HCs and MCI/AD or PSP-Richardson groups were conducted by two-sample t-test. Pearson correlation and linear regression analyses were conducted in a head-to-head comparison among the respective tracers. Clinical associations were also explored by Pearson correlation analysis. Furthermore, voxel-wise analyses were conducted by SPM12; we used the two-sample t-test model of SPM12 for group comparisons. The extent threshold was set to the expected voxels per cluster. For multiple voxel comparisons, family-wise error corrections at cluster levels were applied (*p* < 0.05). All P values are shown in the Figures or their legends.

## DATA AND SOFTWARE AVAILABILITY

Requests for data that support the finding of this study should be directed to the Lead Contact, Makoto Higuchi and will be available upon reasonable request.

## SUPPLEMENTAL INFORMATION

### Method details

Figure S1. Radiosynthesis of ^18^F-PM-PBB3.

Figure S2. Radiometabolites of ^18^F-PM-PBB3 in human subjects.

Figure S3. Brain uptake of ^11^C-PBB3 in human subjects.

Figure S4. List of the VOIs applied to human subjects.

Figure S5. Axial ^18^F-PM-PBB3 PET images of biopsy-confirmed CBD patient.

Figure S6. Distribution of tau lesions in autopsy brain sections of PSP patient.

Figure S7. UV-VIS absorption spectra and photo-isomerization of PM-PBB3 and PBB3.

Figure S8. Non-thresholded ^18^F-PM-PBB3 PET images of a HC, and AD and PSP patients.

Table S1. Metabolite analysis of ^18^F-PM-PBB3 and ^11^C-PBB3 in mouse.

## Notes

### Competing Interest Statement

M-R. Zhang, T. Suhara, M. Higuchi, and H. Shimada hold a patent on compounds related to the present report (JP 5422782/EP 12 884 742.3/CA2894994/HK1208672). All the other authors have no potential conflicts of interest to be disclosed.

### Clinical Trial

UMIN-CTR; number 000030248

